# The impact of rare protein coding genetic variation on adult cognitive function

**DOI:** 10.1101/2022.06.24.22276728

**Authors:** Chia-Yen Chen, Ruoyu Tian, Tian Ge, Max Lam, Gabriela Sanchez-Andrade, Tarjinder Singh, Lea Urpa, Jimmy Z. Liu, Mark Sanderson, Christine Rowley, Holly Ironfield, Terry Fang, Biogen Biobank Team, the SUPER-Finland study, the Northern Finland Intellectual Disability study, Mark Daly, Aarno Palotie, Ellen A. Tsai, Hailiang Huang, Matthew E. Hurles, Sebastian S. Gerety, Todd Lencz, Heiko Runz

## Abstract

Compelling evidence suggests that cognitive function is strongly influenced by genetics. Here, we conduct a large-scale exome study to examine whether rare protein coding variants impact cognitive function in the adult population (N=485,930). We identify eight genes associated with adult cognitive function through rare coding variants with large effects. We demonstrate how the dosage of a single gene, *KDM5B*, may determine the variability of cognitive, behavioral, and molecular traits in mice and humans. We further provide evidence that rare and common variants overlap in association signals and contribute additively to cognitive function. Our findings uncover a contribution of rare coding variants to cognitive function and highlight that the spectrum of cognitive function in the normal adult population is influenced by the action of single genes.

---

Cognitive function in adults, as ascertained either directly via cognitive tests or proxy measures such as educational attainment (EDU), is strongly influenced by genetics and shows substantial genetic correlation with physical and mental health outcomes as well as mortality ^1^. Nearly four thousand cognitive function loci of individually small effect size have been identified through common variant-based genome-wide association studies (GWAS) ^2–6^. GWAS have also demonstrated shared genetic contributions between cognitive function and neurodevelopmental disorders ^7–10^, for which large-scale exome studies in patient cohorts have identified hundreds of underlying genes ^7, 11–13^. However, beyond a proposed deleterious effect of exome-wide rare protein truncating variant (PTV) burden ^14, 15^, no studies have yet systematically interrogated the impact of rare coding variants on cognitive phenotypes in the adult general population.

In order to advance gene discovery for cognitive phenotypes beyond GWAS and gain a deeper insight into the shared genetic components between adult cognitive function and neurodevelopmental disorders, we analyzed exome sequencing and genome-wide genotyping data from 454,787 UK Biobank (UKB) participants with measures of cognitive function. We show that adult cognitive function is strongly influenced by the exome-wide burden of rare protein coding variants and identify and independently replicate eight genes that are individually associated with adult cognitive phenotypes. For one of the cognitive function genes, *KDM5B*, we demonstrate in mice and humans that reduced adult cognitive function at the population-level can be part of a clinical spectrum in which cognitive performance, as reflected by an on average 1.51 less years of schooling in *KDM5B* loss of function carriers, depends on the genetic dose of a single gene. Finally, our study bridges a gap between common and rare disease genetics by demonstrating that adult cognitive function is influenced by additive effects between rare and common variant-based polygenic risk that can be traced to overlapping genomic loci and biological pathways.

## Results

UKB is a prospective cohort study of over 500,000 participants with extensive health and lifestyle data that include genome-wide genotyping and sequencing ^16–22^. We chose to study the genetic basis of three distinct, yet interrelated phenotypes that previous studies have used to approximate adult cognitive function in UKB: educational attainment (EDU), reaction time (RT), and verbal-numerical reasoning (VNR) ^23^. EDU is derived from a survey for years-of-education, which has been shown to be genetically correlated with both adult (R^2^=0.66) and childhood cognitive function (R^2^=0.72) ^5, 24, 25^. RT is based on a digital test that measures processing speed, which is considered a component of general cognitive function ^26, 27^. VNR is a measure of general cognitive function based on questionnaires. EDU and RT were ascertained from all UKB participants at baseline, while VNR was available at baseline from 165,000 participants. We annotated exome sequencing data from 454,787 UKB participants ^19, 28^ for three different classes of coding variation, protein truncating variants (PTVs), missense variants, and synonymous variants ^29, 30^ and identified rare coding variants with minor allele frequency (MAF)<10^-5^ in UKB, in line with previous exome studies on cognitive function-related traits and disorders ^12, 14, 15, 21, 31^. We further annotated all variants by gene intolerance to loss-of-function (LoF) based on pLI (v2.1.1) and missense variants by MPC ^32^ for deleteriousness. In total, we identified 649,321 PTVs, 5,431,793 missense variants, and 3,060,387 synonymous rare variants for analysis.

### Rare variants influence adult cognitive function

We first examined the impact of rare coding variant burden on EDU, RT, and VNR in unrelated UKB participants of European ancestry (EUR; N=321,843; Fig. 1A, Table S1, and S2). We discovered that exome-wide PTV and missense burden have significant deleterious effects on cognitive function (see Methods for measures to correct for population stratification). This was reflected in lower EDU, longer RT, and lower VNR score per variant count (exome-wide PTV burden: *P*=1.95×10^-21^ for EDU, 8.79×10^-19^ for RT, and 6.99×10^-22^ for VNR; missense burden: *P*=5.95×10^-24^ for EDU, 5.95×10^-4^ for RT, and 4.87×10^-12^ for VNR). Consistent with previous exome studies ^12–15, 31^, the most pronounced signals were driven by PTVs and damaging missense variants (MPC>3 and 3≥MPC>2) in LoF intolerant genes (pLI≥0.9) ^33, 34^. Effect sizes of PTV and MPC>3 missense burden in LoF intolerant genes were not significantly different (e.g., for EDU, β=-0.087 with 95% confidence interval [CI]: −0.094 to −0.081 for PTV vs β=-0.069 with 95% CI −0.093 to −0.044 for MPC>3 missense variant), proposing that both classes of variants may similarly impact cognitive function. Synonymous variant burden showed an inverse, albeit small, effect on EDU (exome-wide β=0.0087, *P*=8.59×10^-75^), but not on RT and VNR. In addition to Europeans, we also examined the impact of rare coding variant burden in UKB participants of South Asian (SAS; N=7,086) and African ancestries (AFR; N=6,330) (Table S3, S4, Figs. S1, and S2). However, these analyses were underpowered to replicate the effects of exome-wide rare coding variant burden on cognitive function observed in EUR samples (Methods).

**Fig. 1.**
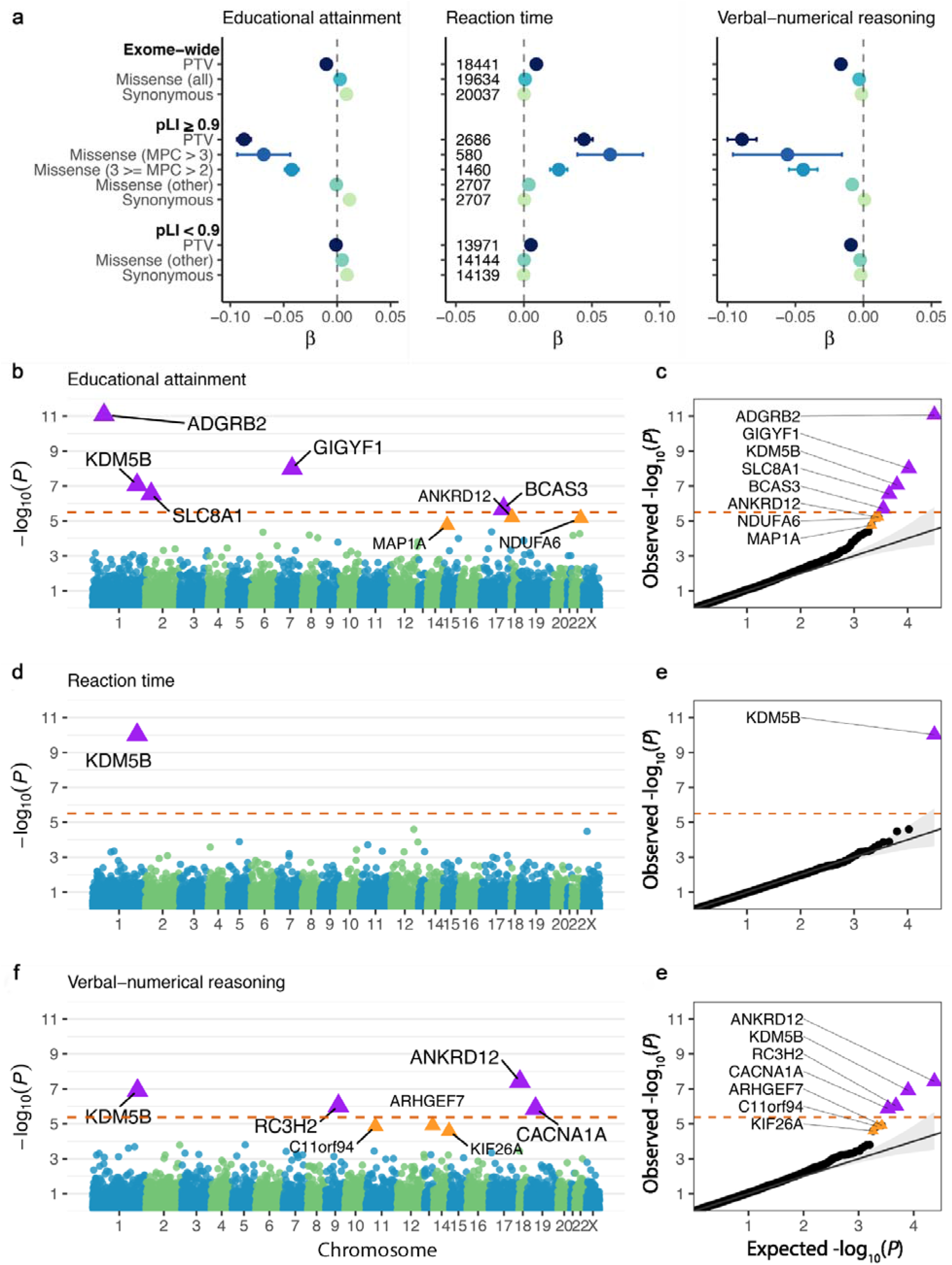
Impact of exome-wide burden of rare protein coding variants and gene discovery based on protein-truncating variant (PTV) burden for educational attainment (EDU), reaction time (RT) and verbal-numerical reasoning (VNR) in European samples in the UK Biobank. **a.** The effects of protein-truncating, missense (stratified by MPC) and synonymous variant burden on EDU, RT and VNR across the exome and stratified by genes intolerant (pLI≥0.9) or tolerant (pLI<0.9) to PTVs. pLI is the probability of being loss-of-function intolerant from the gnomAD database. Missense variants were classified by deleteriousness (MPC) into 3 tiers: MPC>3; 3≥MPC>2; and other missense variants not in the previous two tiers. Number of genes included in each burden was labeled in the middle panel for RT. **b.** Exome-wide gene-based PTV burden association for EDU. The −log10 p-values for each gene were plotted against genomic position (Manhattan plot). Red dashed line indicates the Bonferroni corrected exome-wide significance level per phenotype (*P*<3.17×10^-6^). Purple triangles indicate Bonferroni significant genes. Orange triangles indicate FDR significant genes (FDR *Q*<0.05). **c.** Observed −log10 p-value plotted against expected values (Q-Q plot) for exome-wide gene-based PTV burden association for EDU. Red dashed line indicates Bonferroni corrected exome-wide significance level per phenotype (*P*<3.17×10^-6^ for EDU). **d.** Exome-wide gene-based PTV burden association for RT. Red dashed line indicates the Bonferroni corrected exome-wide significance level per phenotype (*P*<3.16×10^-6^). **e.** Observed −log10 p-value plotted against expected values (Q-Q plot) for exome-wide gene-based PTV burden association for RT. **f.** Exome-wide gene-based PTV burden association for VNR. Red dashed line indicates the Bonferroni corrected exome-wide significance level per phenotype (*P*<4.20×10^-6^). **g.** Observed −log10 p-value plotted against expected values (Q-Q plot) for exome-wide gene-based PTV burden association for VNR.

Following exome-wide burden analyses, we performed gene-based PTV burden tests to identify individual genes associated with EDU, RT, and VNR using two-step whole genome regression implemented in Regenie ^35^. By analyzing 397,434 EUR samples in UKB, we identified eight genes associated with one or more cognitive phenotypes at exome-wide significance after Bonferroni correction (*P*<0.05/15,782=3.17×10^-6^ for EDU, 0.05/15,798=3.16×10^-6^ for RT, and 0.05/11,905=4.20×10^-6^ for VNR; Table 1 and Fig. 1B-G). The Bonferroni corrected-significant cognitive function genes included *KDM5B* (for all 3 phenotypes), *ADGRB2*, *GIGYF1*, *SLC8A1*, *BCAS3* (for EDU), *ANKRD12* (for VNR and for EDU with FDR but not Bonferroni significance)*, RC3H2*, and *CACNA1A* (for VNR). As expected, PTV burden in these 8 genes showed deleterious effects on cognitive function (β=-0.307 to −0.992 for EDU; 0.447 for RT; −0.547 to - 1.126 for VNR) ^14, 15^. We additionally identified 5 putative cognitive function genes at FDR *Q*<5% (*NDUFA6*, *ARHGEF7*, *C11orf94*, *KIF26A*, and *MAP1A*; Table S5).

**Table 1.**
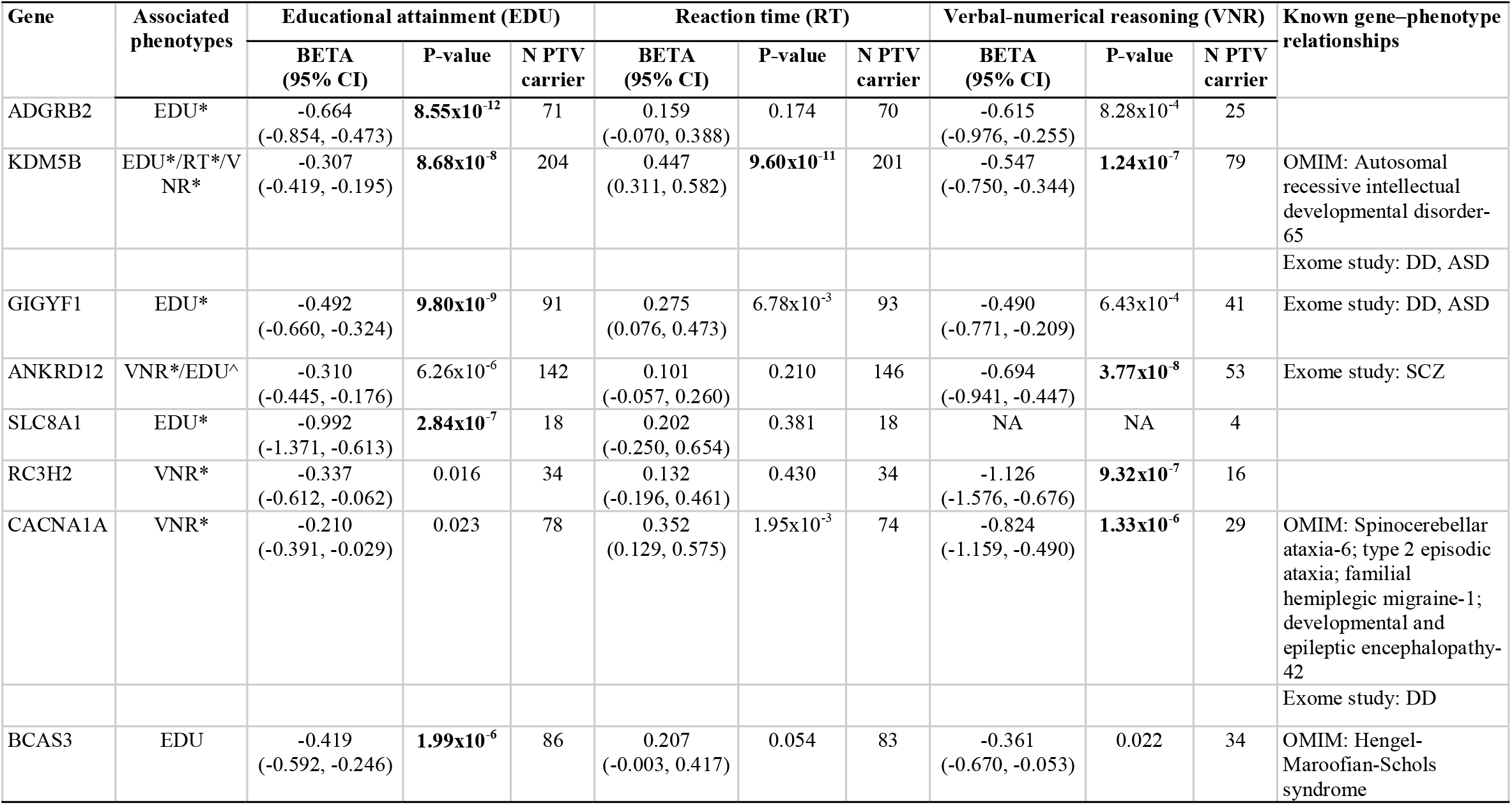
Exome-wide gene-based PTV burden association identified genes for educational attainment (EDU), reaction time (RT) and verbal-numerical reasoning (VNR) in European samples in the UK Biobank. The sample sizes, number of genes tested and λ_GC_for each phenotype are as follows: N_sample_=393,758, N_test_=15,782, and λ_GC_=0.967 for EDU; N_sample_=394,600, N_test_=15,798, and λ_GC_=0.961 for RT; N_sample_=159,026, N_test_=11,905, and λ_GC_=0.959 for VNR. We excluded genes with less than 10 PTV carriers from the analysis. The associated phenotype column indicates the phenotype for each gene with Bonferroni significance (adjusted by N_test_ for each phenotype). *Indicates exome-wide significance after Bonferroni correction across all tests (*P*<0.05/43,485=1.15×10^-6^). ^^^FDR significant for EDU. BETAs are for inverse rank-based normal transferred phenotypes and correspond to SD change in the phenotype. Table sorted by lowest p-value across 3 phenotypes.

We next aimed to replicate our findings from UKB with samples from three independent EUR cohorts: the SUPER-Finland study (N=9,883 psychosis cases), Northern Finland Intellectual Disability (NFID) study (N=1,097 intellectual disability cases and N=11,774 controls) ^36^, and the Mass General Brigham Biobank (MGBB, N=8,389 population cohort), for which both exome sequencing and cognitive function phenotypes were available. We performed association analysis on an aggregated gene set of all 8 cognitive function genes identified in UKB against three cognitive function phenotypes that included developmental disorders/intellectual disability (DD/ID) identified by ICD codes (SUPER-Finland study and NFID study), academic performance (SUPER-Finland study), and educational attainment (SUPER-Finland study and MGBB). Consistent with our findings in UKB, PTV burden was associated with lower educational attainment (β=-0.424, *P*=0.0021), lower academic performance (β=-0.338, *P*=0.0125), and higher risk for DD/ID (odds ratio [OR]=4.812, *P*=8.30×10^-4^) in the SUPER-Finland study (Table S6). The association between the cognitive function gene set and cognitive function in the SUPER-Finland study was conditioned on all samples from this cohort being psychosis cases, which proposes that the observed effects on cognitive function were independent from and on top of potential effects on psychosis. In the NFID study, PTV burden in the cognitive function gene set was also associated with higher risk for DD/ID with OR similar to that estimated in the SUPER-Finland study (OR=4.973, *P*=3.63×10^-5^). The MGBB data showed concordant results in the general population (β=-0.731, *P*=0.5013 for educational attainment). Additional replication analysis including the FDR significant genes (13 genes in total) showed similar results (Table S6). Our replication analyses in three smaller independent EUR cohorts thus independently validate our finding that LoF of the set of genes discovered in UKB influences adult cognitive function.

To systematically assess whether LoF of the eight Bonferroni corrected-significant genes also impacted traits and conditions beyond cognitive function, we conducted PTV burden-based phenome-wide association studies (PheWAS) with 3,150 phenotypes in UKB unrelated EUR samples. Indeed, PheWAS proposed pleiotropy for six of the eight cognitive function genes. For instance, rare PTV burden in *KDM5B* was not only strongly associated with all three cognitive function phenotypes studied (β=-0.307, 95% confidence interval [CI]=-0.419 to −0.195, *P*=8.68×10^-8^ for EDU; β=0.447, 95% CI=0.311 to 0.582, *P*=9.60×10^-11^ for RT; β=-0.547, 95% CI=-0.750 to −0.344, *P*=1.24 x 10^-7^ for VNR), but also showed 16 additional phenome-wide significant associations (Bonferroni corrected p-value threshold <0.05/3150=1.59×10^-5^) related to muscle function (e.g., hand grip strength (right), β=-0.280, *P*=1.02×10^-7^), skeletal phenotypes (e.g., heel bone mineral density T-score, automated (right), *P*=2.93×10^-7^), bipolar disorder (*P*=3.04×10^-7^), and pain medication use (Lyrica/pregabalin, *P*=2.27×10^-10^), among others (Fig. S3 and Table S7). Similarly, PheWAS for *ANKDR12* identified 11 phenome-wide significant additional associations including dysarthria and anarthria (motor disorders with speech deficit; ICD-10 R47.1; *P*=2.28×10^-9^), which hints at a potential mechanism how *ANKRD12* might affect VNR and EDU. Other notable phenome-wide significant associations include type 2 diabetes (and related phenotypes) for *GIGYF1* ^37, 38^ and chlorpromazine (antipsychotic) use and a diagnostic code (R41.8) of impaired cognitive function and awareness for *ADGRB2* (Fig. S4 and Table S7). The substantial pleiotropy indicates these genes do not impact cognitive function in isolation. To provide insights into potential mechanisms, we curated reported OMIM annotations for all 13 genes identified in our exome study (Table 1 and S5) and summarized known and proposed biological functions as well as previously reported associations for these genes in Supplementary Note.

### Cognition and neurodevelopmental genes overlap

Sequencing has identified hundreds of genes underlying developmental disorders (DD) and autism spectrum disorder (ASD) that both disease entities partially share ^12, 13^. As some of the genes we identified are known to cause Mendelian developmental disorders (Table 1), we aimed to elucidate the overall rare genetic variation overlap between adult cognitive function, DD, and ASD through large-scale exome sequencing. We tested whether the rare coding variant burdens in 285 DD-associated genes and 102 ASD-associated genes are associated with adult cognitive function. We observed significant damaging effects of PTV burden in DD and ASD genes on all three cognitive phenotypes analyzed (Fig. 2A and Table S8). Damaging missense variants (MPC>3 and/or 3≥MPC>2) also showed similar deleterious effects.

**Fig. 2.**
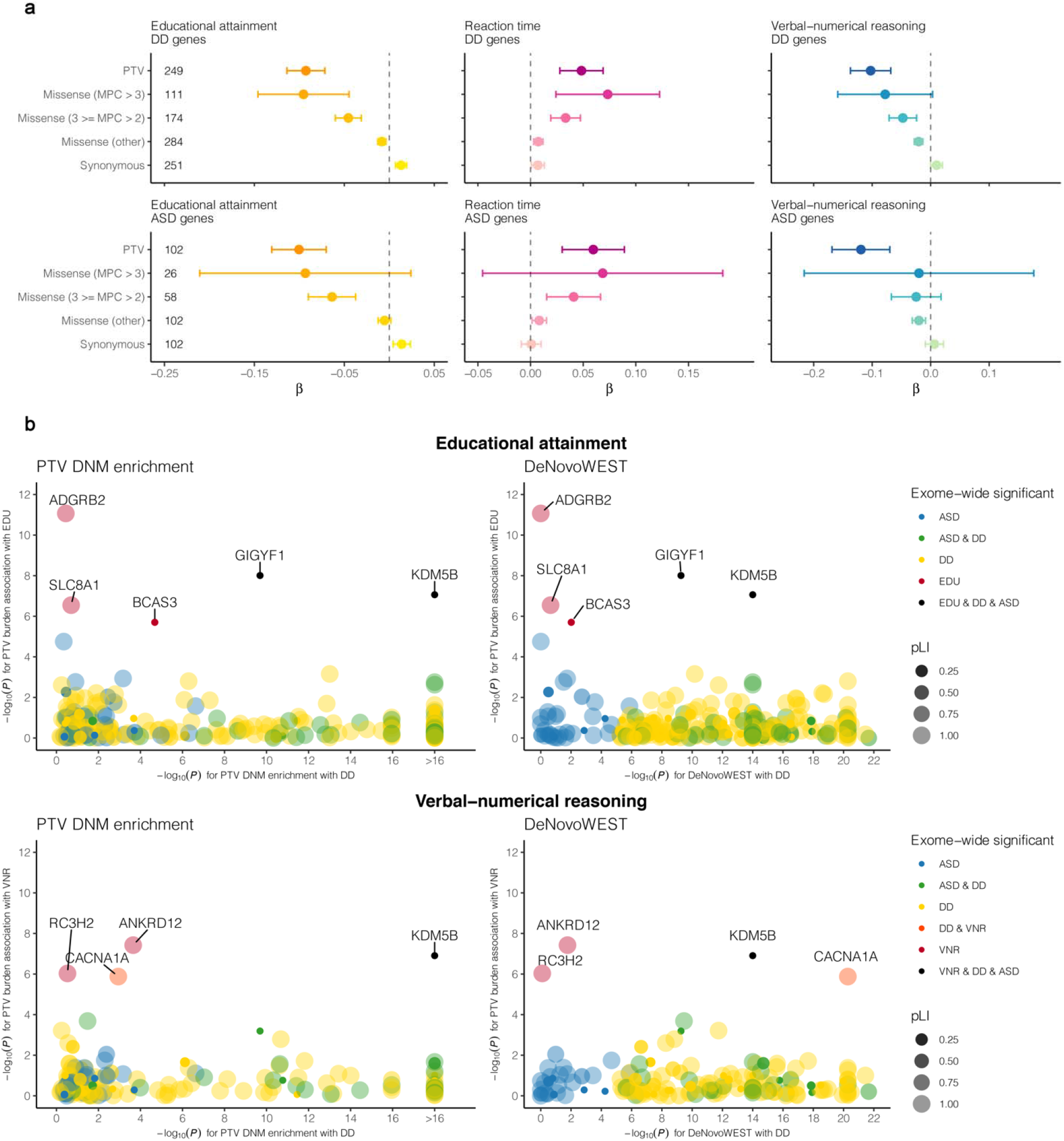
Impact of rare coding variants in developmental disorder (DD) and autism spectrum disorder (ASD) genes on cognitive function. **a.** The effects of protein-truncating, missense (stratified by MPC) and synonymous variant burden in exome sequencing study identified DD genes^13^ and ASD genes^12^ on EDU, RT and VNR. Missense variants were classified by deleteriousness (MPC) into 3 tiers: tier 1 with MPC > 3; tier 2 with 3 ≥ MPC > 2; tier 3 includes all missense variants not in tier 1 or 2. **b.** Comparison between gene-based associations for DD, EDU and VNR (PTV DNM enrichment and DeNovoWEST for DD; rare PTV burden for EDU and VNR). Each dot represents a gene that is identified for DD in Kaplanis et al. 2020 or for EDU or VNR in the current exome analysis. The dots are color-coded according to the phenotypes (DD, ASD, and EDU) that the gene is exome-wide significantly associated with. The size and shade of the dots represent the pLI for the gene. EDU and VNR genes are labeled with gene names.

In order to identify distinct genes linking DD, ASD, and adult cognitive function, we next extracted PTV *de novo* mutation (DNM) enrichment and *de novo* weighted enrichment simulation test (DeNovoWEST) p-values from Kaplanis et al. ^12, 13^ and compared the relative impact of PTV and rare missense variant burden in a combined DD, ASD, EDU, and VNR gene set (Fig. 2B and Table S9). *KDM5B* and *GIGYF1* stood out from these analyses since, interestingly, both genes are LoF tolerant (with pLI scores of 7.91×10^-15^ for *KDM5B* and 3.42×10^-6^ for *GIGYF1*), despite being a cause for DD. *CACNA1A*, a gene encoding for a neuronal calcium channel, was also notable since its association with DD was primarily driven by missense variants (as indicated by a Bonferroni-corrected significant DeNovoWEST), whereas PTV burden was primarily associated with VNR. This is consistent with earlier findings for *CACNA1A*, in which both LoF and GoF mutations may cause neurological diseases with a spectrum of partially overlapping clinical phenotypes ^39–44^. We repeated the above analyses with 2,020 confirmed or probable rare disease genes from the Developmental Disorder Genotype - Phenotype Database (DDG2P) and observed similar results (Fig. S5, Table S8, and S9). Our analyses support that PTVs and missense variants in *KDM5B*, *GIGYF1*, and *CACNA1A* underlie a continuum of conditions with various degrees of cognitive impairment.

### *KDM5B* gene dosage determines clinical phenotype

Homozygous and compound heterozygous mutations in the histone lysine demethylase *KDM5B* are a cause for an autosomal-recessive intellectual developmental disorder with dysmorphic features (OMIM #618109) ^45, 46^. In a heterozygous state, *KDM5B* PTVs were found overrepresented in cases of the Deciphering Developmental Disorders (DDD) study ^7^. To better understand the clinical spectrum of *KDM5B* LoF, we examined the phenotypes documented for individual *KDM5B* PTV carriers in UKB EUR samples (Fig. 3). As expected, cognitive function as approximated by EDU and VNR was on average lower in *KDM5B* PTV carriers (N=204 for EDU and 79 for VNR) than in non-carriers (standardized, residualized phenotype mean=-0.3669 for EDU and −0.5387 for VNR). The degree of cognitive impairment in *KDM5B* carriers was similar between individuals with or without psychiatric, neurodevelopmental, or neurodegenerative diagnoses (Table S10). All individuals carrying heterozygous PTVs that had previously been described as pathogenic or likely pathogenic for intellectual developmental disorder in ClinVar showed reduced EDU and VNR. However, the degree of cognitive impairment did not seem to be more or less pronounced for ClinVar listed versus novel *KDM5B* PTVs. Moreover, none of three UKB participants heterozygous for the pathogenic p.Arg299Ter variant (rs1558498928; NM_006618.5:c.895C>T) had diagnostic records of intellectual developmental disorder. We identified 35 *KDM5B* PTV carriers who had been diagnosed with psychiatric disorders (schizophrenia, depression, bipolar disorder, substance use, anxiety, and stress disorders), epilepsy or Parkinson’s disease based on ICD-10 (International Statistical Classification of Diseases and Related Health Problems, 10^th^ revision) codes (enriched for disease cases compared with EUR non-*KDM5B* PTV carriers; *P*=0.0005). However, EDU was impaired to a similar extent in *KDM5B* PTV carriers with and without such comorbidities, while VNR was reduced even further (*P*=0.032). We found similar evidence for low cognitive function among *CACNA1A* PTV carriers (Fig. S6 and Table S11). Among a total of 78 PTV carriers for *CACNA1A*, we identified 7 carriers for 5 pathogenic variants annotated by ClinVar but none of these carriers have any in-patient diagnostic record that would suggest Mendelian developmental disorders. We also identified 5 depression or epilepsy patients among the PTV carriers. Furthermore, we showed average lower cognitive function in *CACNA1A* PTV carriers compared with non-carriers, which suggests a potentially milder form of cognitive impairment caused by PTVs in *CACNA1A* (Fig. S6 and Table S11).

**Fig. 3.**
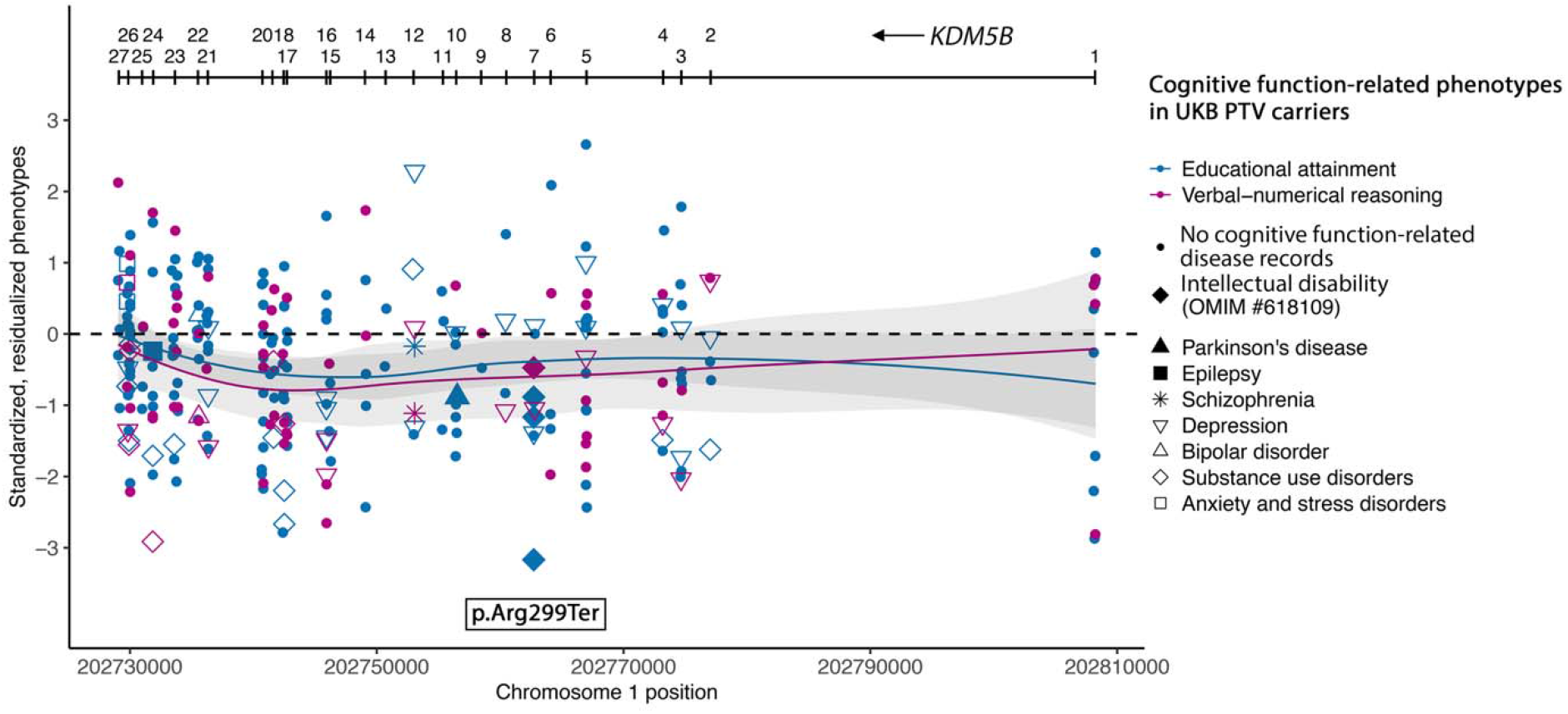
Phenotypic characterization and gene expression of *KDM5B* in human. Distribution of educational attainment and verbal-numerical reasoning score for *KDM5B* PTV carriers in UKB. ClinVar likely pathogenic variants for intellectual developmental disorder (OMIM #618109) were annotated. Samples with inpatient ICD-10 (International Classification of Diseases version-10) records of psychiatric (schizophrenia, bipolar disorder, depression, substance use disorder and/or anxiety and stress disorders), neurodegenerative and neurodevelopmental disorders were annotated. Phenotypes were residualized by sex, age, age^2^, sex by age, sex by age^2^, top 20 PCs and recruitment centers and were inverse rank-based normal transformed.

Based on our findings in UKB and *KDM5B’*s known role in disease, we hypothesized that lower EDU and VNR in heterozygous *KDM5B* PTVs carriers may be explained by a *KDM5B* gene dosage effect where adult heterozygous PTV carriers show an attenuated phenotype relative to individuals with *KDM5B* LoF mutations on both alleles with childhood-onset intellectual disability. To further examine the relationship between *KDM5B* gene dosage and cognitive function, we conducted a series of cognitive and behavioral tests in a previously reported *Kdm5b* mouse model ^45^. Relative to wildtype siblings, both heterozygous and homozygous *Kdm5b* mutant mice showed cognitive, behavioral, and skeletal phenotypes, with respective phenotypes consistent with an additive effect of *Kdm5b* LoF (Fig. 4A). Specifically, mutant mice showed deficits in spatial memory (Barnes maze), reduced novel object recognition, as well as evidence for behavioral abnormalities such as increased anxiety (light-dark box). Furthermore, skeletal abnormalities observed in homozygous knockout mice, such as changes in craniofacial dimensions or transitional vertebrae, were also present in heterozygous *Kdm5b* mice, yet at an intermediate severity and/or frequency (Fig. S7). An additive effect of *Kdm5b* LoF was further supported by *Kdm5b* mRNA levels in whole brain of embryonic, as well as frontal cortex (FrC), hippocampus (HIPP), and cerebellum (CR) of adult heterozygous mice being at intermediate levels of that of wildtype and homozygous mutant mice (Fig. 4B). Consistently, 92% of the 723 genes identified by RNA sequencing (RNAseq) as differentially expressed (union of DE genes in both mutant genotypes, FDR Q<0.1) in *Kdm5b* mutant mouse brain showed the same directionality of change in both homozygous and heterozygous mice, with a globally smaller effect size in the heterozygous state than in the homozygous state (Fig. 4C and Fig. S8). We also showed that brain expression of *Kdm5b* in mice is higher during embryonic stages than in adult tissues (Fig. 4B). Notably, *KDM5B* mRNA levels in brain also vary across the human lifespan ^47^, with highest expression prenatally (*P*=8.06×10^-165^ relative to postnatally) and a decline at later developmental stages (Fig. S9, Tables S12, and S13). Consistently, more genes are differentially expressed in embryonic *Kdm5b* mutant mice than in adults, with a strong enrichment for genes with roles in brain development, synapse function and brain structure (Fig. 4C, Fig. S10, Table S14, and S15). In summary, our data from both mice and humans provide strong evidence that *KDM5B* LoF modulates cognition, behavior, and skeletal phenotypes as well as brain mRNA expression to an extent that directly correlates with *KDM5B* genetic dose.

**Fig. 4.**
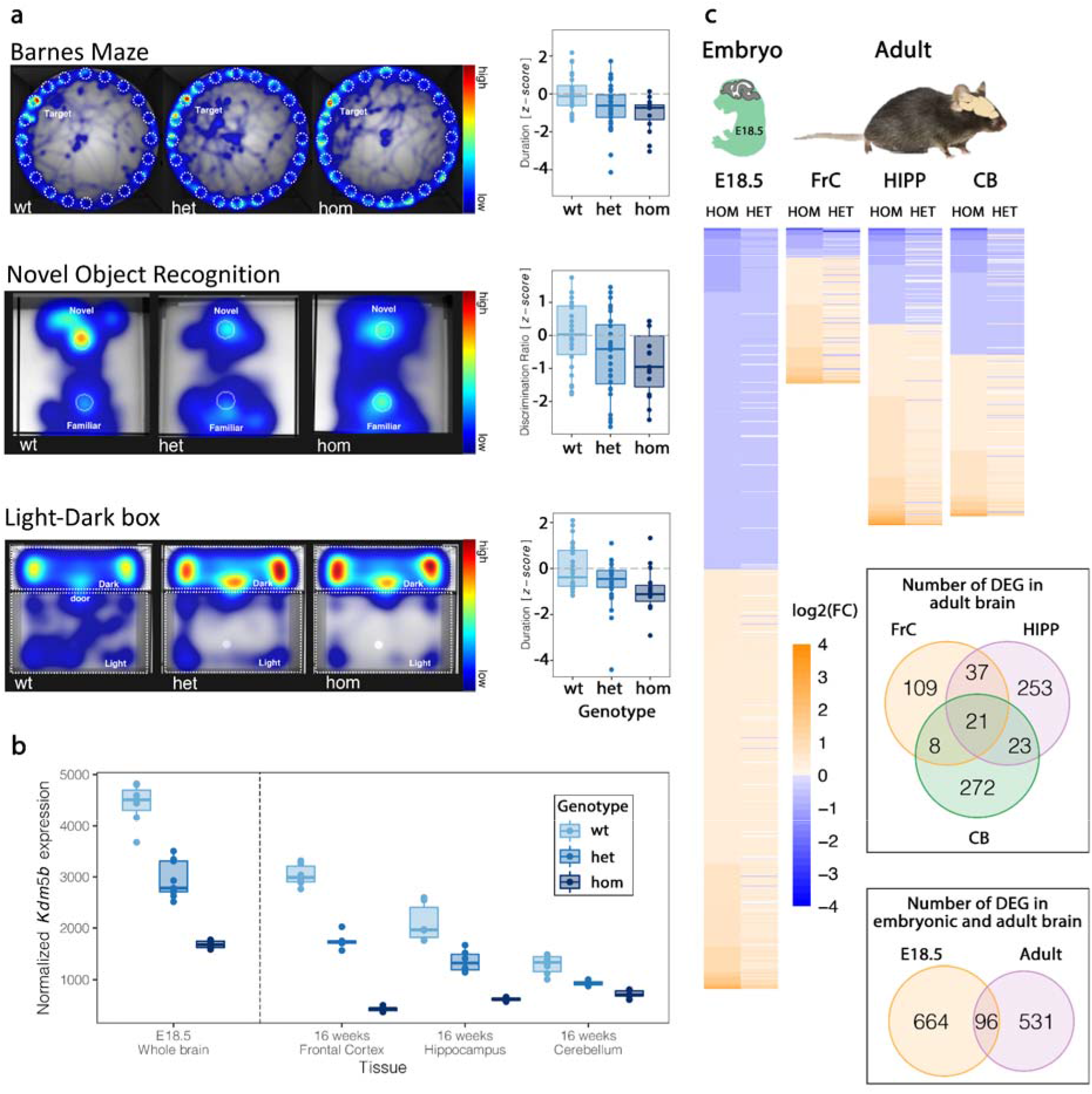
*Kdm5b* LoF alleles display a gene dosage effect on behavioral, cognitive, and molecular phenotypes in mice. **a.** Mice carrying *Kdm5b* LoF alleles show a dose-dependent decrease in spatial memory performance (Barnes Maze; additive genotype effect with *P*=0.042; *Kdm5b^+/-^* N=34, *P*=0.050 and *Kdm5b-/-* N=15, *P*=0.001 spent less time around goalbox [GB], compared to wildtype control animals, N=24), decrease in object recognition memory performance (New Object Recognition; additive genotype effect with *P*=0.0087; *Kdm5b^+/-^*N=32, *P*=0.038 and *Kdm5b^-/-^* N=15, p=0.011 had reduced discrimination compared to wildtypes, N=26) and increase in anxiety related behavior (Light-Dark box; additive genotype effect with *P*=0.0083, *Kdm5b^+/-^* N=15, *P*=0.025 and *Kdm5b^-/-^* N=34, *P*=0.0037 spending more time in dark compared to wildtypes). Heatmaps showed relative time spent around various arenas during the trial period of each assay, as a composite of all mice of the same genotype (Barnes Maze and Light-Dark box) or the trace for a single representative animal (Novel Object Recognition). *Kdm5b^+/-^* (het) and *Kdm5b^-/-^* (hom) mice spend less time around GB (Barnes Maze), show reduced discrimination of the novel object (Novel Object Recognition) and spend more time in the dark zone (Light-Dark box) compared with wildtypes. **b.** Normalized read counts of *Kdm5b* gene expression in wildtype, *Kdm5b^+/-^* and *Kdm5b^-/-^* mice across embryonic and adult tissues as indicated. **c.** Heatmap of expression changes (log2 fold change) in differentially expressed genes in *Kdm5b^+/-^* (het) and *Kdm5b^-/-^* (hom) mice across embryonic and adult tissues as indicated. There is a strong correlation between direction of change in expression in both mutant genotypes. Venn diagrams showed the overlap of differentially expressed genes (DEGs) in both *Kdm5b^+/-^*and *Kdm5b^-/-^* mice across tissues and stages.

### Rare and common variant signals intersect

We further tested whether genes identified through our PTV burden analysis in UKB overlap with genetic loci identified in previous common variant-based educational attainment ^5^ and cognitive function GWAS ^48^. Indeed, we identified overlapping signals between an educational attainment GWAS locus on chromosome 1 and *ADGRB2* which we had found associated with EDU in our PTV burden analysis. Notably, the PTVs for which carriers showed lower than average EDU and VNR and the GWAS top associated SNPs were in close genomic proximity, prioritizing *ADGRB2* as the most likely underlying causal gene of the GWAS association signal over other genes located in the nearby region (Fig. S11). We also identified overlapping signals between a locus on chromosome 22 in cognitive function GWAS and *NDUFA6*, an FDR-significant cognitive function gene associated with EDU, prioritizing *NDUFA6* over other genes at this GWAS locus (Fig. S12).

To further characterize the overlap between rare coding variant association and common variant associations with cognitive function, we utilized UKB exomes to calculate rare coding variant burdens for genes identified in cognitive function phenotype GWAS (see Methods). PTV burden in educational attainment GWAS genes showed significant effects on all three cognitive phenotypes (β=-0.023 and *P*=3.69×10^-7^ for EDU; β=0.017 and *P*=6.38×10^-5^ for RT; β=-0.033 and *P*=4.05×10^-6^ for VNR), while PTV burden in cognitive function GWAS and schizophrenia GWAS genes showed significant effects on EDU (Figs. S13-S15 and Table S16). Significant effects were also observed for missense variants. Missense burden (3≥MPC>2) in cognitive function GWAS genes showed significant effects on VNR (β=-0.060 and *P*=3.01×10^-4^) and EDU (β=-0.045 and *P*=1.00×10^-5^; Figs. S13, S15, and Table S16). In addition, missense burden (3≥MPC>2) in schizophrenia GWAS genes also showed significant effects on EDU (β=-0.033 and *P*=7.57×10^-6^; Figs. S13, S15, and Table S16).

GWAS have identified biological pathways of potential relevance to cognitive function ^2,3^. To further explore biological mechanisms through which rare variants might impact cognitive function, we performed PTV burden analysis for 13,011 gene sets from the Molecular Signatures Database (MSigDB v7.2; C2 canonical pathways and C5 gene ontology pathways) in UKB (Fig. S16 and Table S17). We identified 182, 66 and 56 Bonferroni corrected significant gene sets for EDU, RT and VNR, respectively. The most significant gene sets are involved in synaptic function, neurogenesis, neuronal differentiation, and neuronal development. These signatures highly overlap with those from cognitive function GWAS^2^ (such as neurogenesis), suggesting that rare and common variants modulate cognitive function through similar mechanisms. Further analyses showed that the burden of PTV and damaging missense variants in genes with brain specific expression impact cognitive function more strongly than in genes primarily expressed in other tissues (Fig. S17 and Table S18), which is also consistent with previous GWAS findings^2, 3, 5^.

Finally, we explored the relationship between rare coding variants and common variant-based polygenic risk on cognitive function. We calculated polygenic risk scores (PRS) in unrelated EUR samples in UKB using imputed genome-wide genotype data and SNP weights based on cognitive function GWAS (from Cognitive Genomics Consortium [COGENT] excluding UKB samples) ^4^ using PRS-CS ^49^, where higher PRS reflects genetic liability of *increased* cognitive function. We tested for the joint effects of PRS and carrier status for PTVs and/or MPC>2 damaging missense variants in LoF intolerant genes (pLI ≥ 0.9). These analyses showed that the effect of PRS and rare coding variants is additive (PRS interaction test *P*=0.27 for PTV and *P*=0.21 for damaging missense for EDU; Fig. 5 and Table S19). For EDU, conditional effects (β) of PRS, PTV carrier status, and damaging missense carrier status were 0.116, −0.095, and −0.053, while the adjusted partial R^2^ were 0.013, 0.0015, and 0.0005, respectively (*P*=8.96×10^-949^ for PRS, 7.33×10^-109^ for PTV and 6.76×10^-38^ for missense variants). Our results strongly suggest that the prediction of genetic liabilities of cognitive function through common variant-based PRS can be further refined by integrating rare coding alleles. We repeated these analyses for VNR and reached similar conclusions (Fig. S18 and Table S20).

**Fig. 5.**
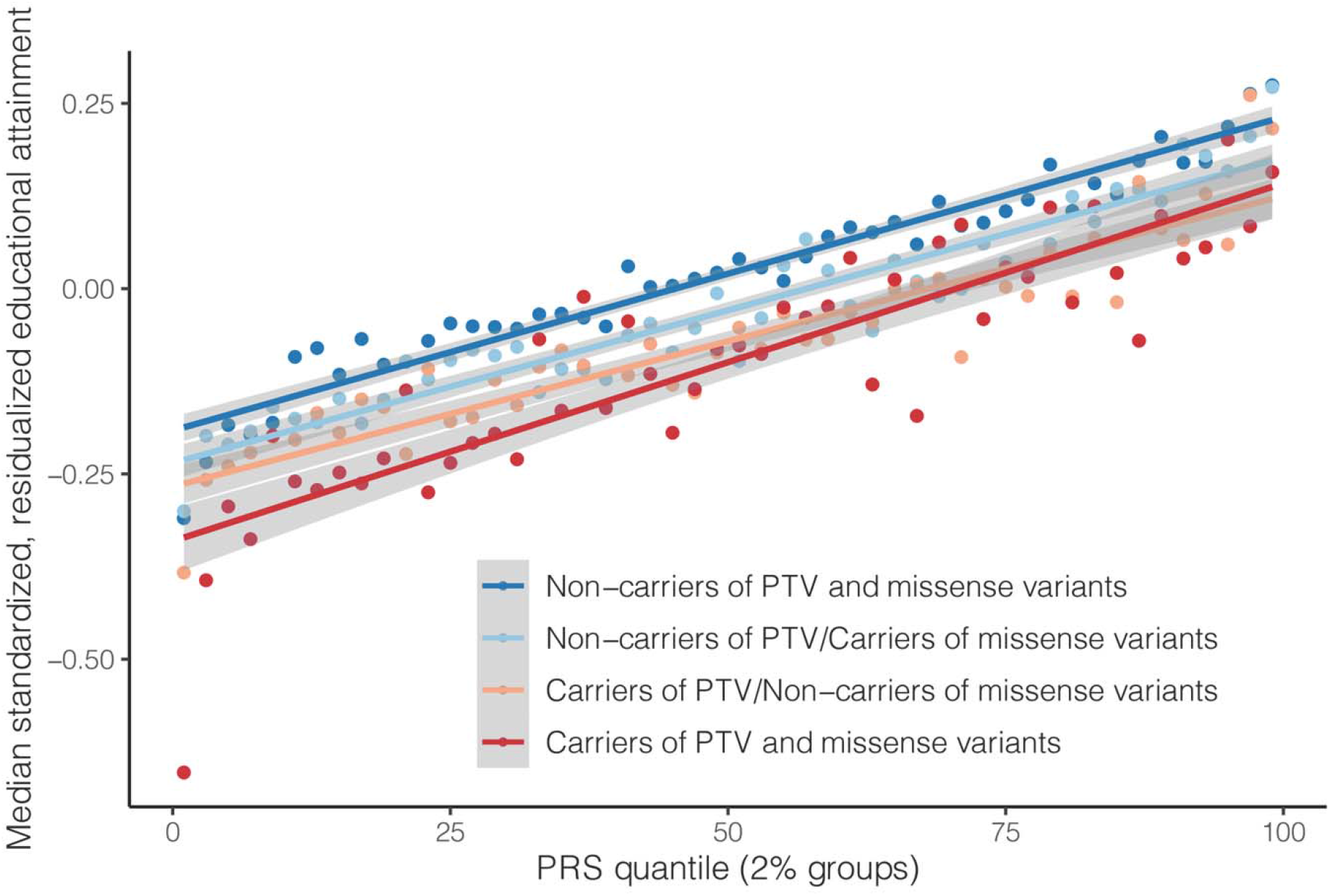
Contribution of common variants and rare coding variants to educational attainment (EDU). The impact of cognitive function polygenic score (PRS) and carrier status of PTV and/or damaging missense variants (MPC>2) in LoF intolerant genes (pLI>0.9) on EDU. Educational attainment was residualized by sex, age, age^2^, sex by age, sex by age^2^, top 20 PCs and recruitment centers and inverse rank-based normal transformed. Median of educational attainment was calculated for individuals stratified by PRS quantiles (in 2% groups) and PTV and/or damaging missense variant carrier status.

## Discussion

Here, we present the first large-scale exome sequencing study on cognitive function and educational attainment in the adult general population. Our findings support previous evidence that an increased exome-wide burden of rare PTVs is associated with lower cognitive function ^14, 15^ and extend this observation to deleterious missense variants. The large number of exome-sequenced participants in UKB allowed us to identify eight genes associated with measures of cognitive function at Bonferroni-corrected exome-wide significance. A relevance for these genes for cognitive function is supported by three independent European cohorts. Notably, several of the cognitive function genes have established roles in neurodevelopmental disorders, suggesting that at least in some cases reduced cognitive function in participants of population cohorts such as UKB may be due to defects in single genes, thus extending the spectrum of neurodevelopmental conditions to the normal adult population.

Our study is a natural extension of previous GWAS on cognitive function and educational attainment that have primarily investigated the role of common genetic variation on cognitive traits ^2–6^. While highly successful in identifying associated loci, applying the GWAS approach to cognitive function has also received substantial criticism. Typically, common variant associations observed in GWAS are susceptible to systematic biases from ascertainment, such as ancestry, geography, and environmental or cultural differences between subpopulations ^25, 50^. Cognitive function is difficult to assess in isolation since there is substantial genetic and epidemiological overlap with other traits. For instance, EDU is not only reflective of childhood and adult IQ, but also strongly correlates with other traits including income, parental age at birth, alcohol dependence, or neuroticism ^25^. Nevertheless, we are confident that for several reasons the results of our exome-study are less susceptible to such biases than traditional GWAS. First, we analyzed three distinct phenotypes (EDU, RT, and VNR) that each capture different aspects of cognitive function. The consistency of exome-wide, gene set-level, and gene-level associations across EDU, RT, and VNR, which translate also to independent exome-sequenced cohorts, increases the confidence that our findings are indeed biologically relevant for cognitive function. Second, five of the eight cognitive function genes (*KDM5B* ^12, 13^ [OMIM #618109], *ANKRD12* ^31^*, CACNA1A* [OMIM #138086, 108500, 617106]*, GIGYF1* ^12, 13^, and *BCAS3* [OMIM #619641]) are either established Mendelian developmental disorder genes or have also been identified in previous exome studies on schizophrenia ^31, 51^, DD ^13^ or ASD ^12^. A sixth gene, *ADGRB2*, resides within an educational attainment GWAS locus ^5^ and on top of its association with EDU was identified through our PTV burden-based PheWAS as also associated with ICD-10 code R41.8 reflecting signs of cognitive dysfunction. A biological relevance for the genes discovered in this study across multiple relevant conditions is consistent with the well-established tight genetic relationships between cognitive traits and diseases. Third, coding variants as analyzed here can sometimes yield clues to a gene’s biological mechanisms. For instance, we found that variants in *ANKRD12*, a gene that met Bonferroni-corrected significance for both EDU and VNR but has not previously been linked to DD, are also associated with dysarthria/anarthria, myasthenia gravis and disorders of calcium metabolism. This suggests that cognitive dysfunction in individuals with *ANKRD12* LoF is accompanied by imbalances in motor coordination or muscle function and might be part of a yet undescribed genetic syndrome. Likewise, variants in *CACNA1A*, which encodes for the neuronal calcium channel P/Q, underlie a broad spectrum of autosomal dominant conditions ranging from early developmental delay with epileptic encephalopathy over spinocerebellar ataxia 6 to paroxysmal disorders like familial hemiplegic migraine through both GoF and LoF mechanisms ^44^. The substantial increase in association signal between *CACNA1A* missense variants, but not PTV burden, and DD indicates that reduced VNR in rare variant carriers might result from alterations in channel function rather than an absence of the *CACNA1A* protein.

A particularly intriguing example for how diseases with cognitive impairment and adult cognitive function intersect is *KDM5B*, a histone lysine demethylase with roles in neuronal differentiation ^45, 52–54^ that we interrogated further in humans and mice. Biallelic mutations in *KDM5B* cause autosomal recessive intellectual developmental disorder ^45, 46, 53^, while heterozygous PTVs have been linked to severe DD ^7, 13, 45^ and ASD ^12^ with presumed incomplete penetrance ^45, 55^. However, since previous studies focused on patients, the relevance of *KDM5B* LoF on adult cognitive function in the general population has not yet been appreciated. Unlike most other DD genes ^13^, *KDM5B* is LoF-tolerant, leading to a relatively high PTV-carrier rate of ∼1:1,900 participants in UKB. Since UKB participants tend to be healthier and more educated than the general UK population ^56, 57^, it can be expected that *KDM5B* PTV carrier rates in the general European population are even higher. With this, our results strongly propose a gene-dosage effect for *KDM5B*, where damaging variant carriers at the more severe end of a cognitive function spectrum show a biallelic, near complete loss of *KDM5B* function and will be observed in patient cohorts, whereas damaging variant carriers at the less severe end of the phenotypic spectrum will present with only moderate cognitive impairment that overlaps with the spectrum of cognitive function in the normal population. Notably, beyond cognitive function, *KDM5B* was also associated with muscle strength, bone density, growth hormone levels, and bipolar disorder, among other phenotypes, in our phenome-wide PTV burden analysis. This pleiotropy partially overlaps with the dose-dependent cognitive, behavioral, and skeletal symptoms in our *Kdm5b* mouse model ^45^ as well as *KDM5B* patients ^46^. Genes with a similar dosage sensitivity like described here for *KDM5B* are considered ideal drug targets since the degree of genetic impairment may guide the development of gene-directed therapeutic interventions ^58, 59^. KDM5B is already an established drug target with molecules inhibiting its enzymatic activity in preclinical development for cancer ^60^. It could be interesting to explore whether activators exist that might improve cognitive phenotypes ^61, 62^.

A particular strength of exome studies is that genes and variants identified through rare variant tests tend to exhibit much larger effect sizes than common variants identified in GWAS. For example, heterozygous carriers of *KDM5B* PTVs show on average 1.51 years-of-schooling less than non-carriers. In contrast, lead SNPs in the most recent EDU GWAS based on three million individuals only show a median 1.4 week increase in schooling per allele (with the 5th and 95th percentiles of the estimated effect being 0.9 and 3.5 weeks) ^6^. This demonstrates that exome studies may uncover substantially stronger effects and complement GWAS to describe the genetic architecture of cognitive function more comprehensively. This is further supported in the case of *ADGRB2* and *NDUFA6*, which our results propose as the most likely causal genes underlying GWAS peaks in EDU and cognitive function GWAS (Fig. S11 and S12) ^5^.

With both exome sequencing and genome-wide genotype data available in UKB, we were able to explore the relative contribution of common variant-based polygenic risk and rare coding variant burden to cognitive function. Our results provide evidence that PTVs and damaging missense variants affect EDU and VNR additively to PRS and thus propose that the accuracy of genetic prediction can be further improved by combining PRS and rare coding variant burden. Similar findings were reported previously for other common complex phenotypes including breast cancer, colon cancer or coronary artery disease ^51, 63–65^. Consistent with other diseases, due to their rarity, the phenotypic variance for EDU and VNR explained by damaging coding variants is much smaller than that explained by PRS. However, the rare coding variant burden provides orthogonal predictive power that is not relying on external training data (e.g., GWAS), which is required for PRS, and is thus less susceptible to population stratification ^66^.

Future studies will be needed to better understand the biological basis of how the genes and variants reported here impact cognitive traits and related diseases. Moreover, our findings do not imply direct applications in clinical practice, such as for prenatal genetic screening ^67, 68^ and should be interpreted with similar caution as reported for GWAS ^6^. Further work will also be needed to assess how well results from our study can be extrapolated to other Europeans as well as other ethnicities. Nevertheless, our results provide a starting point towards expanding our knowledge on how rare genetic variants impact cognitive function at the population level and support a convergence of rare and common genetic variation jointly contributing to the spectrum of cognitive traits and diseases.

## Methods

### Cognitive function phenotypes in UK Biobank

The UK Biobank (UKB) is a prospective cohort study of the UK population with over 500,000 participants ^16^. Participants were aged between 40 to 69 years at recruitment in 2006-2010 and provided extensive phenotype data, including surveys on baseline characteristics and health outcomes, specific questionnaires and assessments, inpatient hospital records, physical measures and biomarkers ^17^.

We extracted three cognitive function phenotypes for our analysis: educational attainment (EDU), reaction time (RT) and verbal-numerical reasoning (VNR) ^23^. Educational attainment is based on a baseline survey for years of schooling that reflects both cognitive function and non-cognitive components ^24^. We extracted UKB data field 6138 “Qualifications” collected at baseline as a measure of educational attainment ^5^. The educational attainment survey is a multiple-choice question with eight choices, including 7 categories of different qualifications and an option for “Prefer not to answer”. We mapped the seven categories to years of schooling using the International Standard Classification of Education (ISCED) scale: none of the above (no qualifications) = 7 years of education; CSEs or equivalent = 10 years; O levels/GCSEs or equivalent = 10 years; A levels/AS levels or equivalent = 13 years; other professional qualification = 15 years; NVQ or HNC or equivalent = 19 years; college or university degree = 20 years of education ^5^. Educational attainment for those who selected “Prefer not to answer” was treated as missing and excluded from the analysis.

Reaction time is a measure of processing speed, which is a component of general cognitive function ^2, 4, 26, 27^, implemented as a digital assessment at baseline. We extracted the UKB data field 20023 “Mean time to correctly identify matches” from the “Snap” game. In the “Snap” game, the participants were presented with pairs of matched or mismatched cards on the computer screen. If the two cards were matched, participants were to push a button box as quickly as possible. The game includes first five rounds of practice and then seven rounds of the actual tests, among which four rounds present matched pairs of cards. “Mean time to correctly identify matches” is the mean time to correctly identify the four rounds of matched cards (in milliseconds) ^2^.

Verbal-numerical reasoning is measured by a structured questionnaire focusing on assessing crystalized and fluid intelligence in both verbal and numerical aspects. We extracted the VNR score from UKB data fields 20016 “Fluid intelligence score”. The “fluid intelligence” questionnaire contains 13 multiple-choice questions that assess verbal and numerical problem solving. The score is the total number of correctly answered questions in two minutes. Note that only a subset of 165,453 UKB participants completed the VNR assessment at baseline, while EDU (N = 497,844 at baseline collection) and RT (N = 496,660 at baseline collection) data were collected for almost the entire UKB cohort.

By examining three cognitive phenotypes, we aimed at capturing different aspects of cognitive function for a more comprehensive discovery and to facilitate a dissection of the genetics of cognitive function. For association analyses in UKB samples, we inverse rank-based normal transformed the phenotypes. We note that while higher EDU and VNR score indicate better cognitive function, longer RT represents worse cognitive function.

### The UK Biobank whole-exome sequencing data

Whole-exome sequencing data from UK Biobank participants was generated by the Regeneron Genetics Center (RGC) on behalf of the UKB Exome Sequencing Consortium, which is a collaboration between AbbVie, Alnylam Pharmaceuticals, AstraZeneca, Biogen, Bristol-Myers Squibb, Pfizer, Regeneron, and Takeda.

Whole-exome sequencing data production was conducted at the RGC as described in VanHout et al^28^. Briefly, whole-exome sequencing was done on an Illumina NovaSeq 6000 platform using xGen exome capture kits. Sequencing reads were aligned to the GRCh38 reference genome using BWA-mem ^69^. SNVs and INDELs were called by first generating gVCF files using WeCall variant caller (Genomics Plc.; https://github.com/Genomicsplc/wecall) and then joint called by GLnexus joint genotyping tool ^70^. The joint called project-level VCF (pVCF) was then filtered by the RGC quality control (QC) pipeline (the “Goldilocks” set). As of November 2020, we obtained QC passed whole-exome sequencing data from 454,787 UKB participants. UKB has released these data publicly to approved researchers via their Research Analysis Platform. For instructions on how to access these data, see https://www.ukbiobank.ac.uk/enable-your-research/research-analysis-platform.

We annotated variants by Variant Effect Predictor (VEP) v96 ^29^ with genome build GRCh38. Variants annotated as stop-gained, splice site disruptive and frameshift variants were further assessed by Loss-Of-Function Transcript Effect Estimator (LOFTEE) ^30^, a VEP plugin. LOFTEE implements a set of filters to remove variants that are unlikely to be disruptive. Those variants labeled as “low-confidence” were filtered out, while variants labeled as “high-confidence” were kept. Variants annotated as missense variants were annotated for deleteriousness by MPC score^32^. We also annotated variants based on gene intolerance to loss-of-function using pLI (probability of being loss-of-function intolerant; v2.1.1) ^30, 33^. All predicted variants were mapped to GENCODE ^71^ canonical transcripts. In total, we identified 649,321 predicted rare protein truncating variants (PTVs), 5,431,793 missense variants and 3,060,387 synonymous variants with minor allele frequency (MAF) < 1.0×10^-5^.

We note that in previous smaller-scale exome studies, educational attainment, intellectual disability, and psychiatric disorders (e.g., autism, attention-deficit/hyperactivity disorder, schizophrenia) were associated with the burden of ultra-rare variants at exome-wide level. In these studies ultra-rare variants were defined as variants observed in fewer than 1 in 74,839 individuals (allele frequency less than 1.34×10^-5^) or 1 in 201,176 individuals (allele frequency less than 2.49×10^-6^) ^14, 15^. A recent large-scale exome study for schizophrenia also adopted a minor allele count cut-off of less than 5 alleles in 24,248 cases and 97,322 population controls, which corresponds to a MAF cut-off of 2.06×10^-5^, and identified novel schizophrenia associated genes ^31^. In line with these earlier exome studies, our current study focuses on coding variants with MAF less than 1.0×10^-5^ in UKB to enrich for pathogenic variants.

### The UK Biobank genome-wide genotype data

We used imputed genotype data provided by UKB with additional QC filtering. Genome-wide genotyping had been performed for all UK Biobank participants and imputed using the Haplotype Reference Consortium (HRC) ^72^ and UK10K ^73^ plus 1000 Genomes Project reference panels ^74^, resulting in a total of more than 90 million variants. We performed QC on the genotyping data by filtering out variants with imputation quality INFO score < 0.8 and variants with MAF less than 0.01 by PLINK2 ^75^. We filtered out 1,804 individuals whose reported gender differed from genetic gender, individuals showing sex chromosome aneuploidies, as well as 133 individuals who had withdrawn from the UK Biobank (as of August 24, 2020).

To identify UK Biobank samples from different ancestral populations, we performed population assignment based on population structure using principal component analysis (PCA) with 1000 Genomes Project (1KG) reference samples (N sample = 2,504) from 5 major population groups: East Asian [EAS], European [EUR], African [AFR], American [AMR] and South Asian [SAS]. We first performed quality control on the 1KG genotype data by retaining only the SNPs on autosomes with minor allele frequency (MAF) > 1% and removed SNPs located in known long-range LD regions (chr6:25-35Mb; chr8:7-13Mb). We also removed 1 sample from each pair of related samples (greater than second degree) in 1KG. We merged the UK biobank imputed genotype data that was filtered to imputation quality INFO > 0.8 and MAF > 1% with the 1KG genotype data and performed LD-pruning at R^2^ = 0.2 with a 500 kb window. We then computed principal components (PCs) using the LD-pruned SNPs in 1KG sample and derived projected PCs of UK Biobank samples using the SNP-wise PC loadings from 1KG samples. Using the 5 major population labels of 1KG samples as the reference, we trained a random forest model with top 6 PCs to classify UK Biobank samples into 1KG population groups. We assigned UK Biobank samples into one of the 5 populations defined with 1KG reference based on a predicted probability for a specific population group > 0.8. We identified 1,609 EAS samples, 458,197 EUR samples, 8,406 AFR samples, 9,224 SAS samples, 1,085 AMR samples and 8,874 samples without explicit population assignment. Note that the final analytical sample sizes for each population are smaller due to the fact that exome sequencing was available for only 454,787 UKB samples, while population assignment was done for all UKB samples with genome-wide genotyping data. Due to the small sample sizes, we did not further analyze samples in EAS and AMR groups. We also did not further analyze samples without an explicit population assignment. After initial population assignment, we performed three rounds of within-population PCA for AFR, EUR and SAS samples to identify remaining population outliers, each time removing samples with any of the top 10 PCs that was more than 5 standard deviations (SD) away from the sample average. We used the in-sample PCs derived after outlier removal for each population in subsequent analyses.

### Gene set-based rare coding variant burden test

#### Analysis overview

To estimate the association between cognitive function phenotypes (EDU, RT and VNR) and gene set-based rare coding variant burdens, we inverse rank-based normal transformed (IRNT) the phenotypes and fitted linear regression in unrelated UKB samples within samples from the same population group (as described in the previous section on population assignment). To minimize potential population stratification and confounding, we adjusted for sex, age, age^2^, sex by age interaction, sex by age^2^ interaction, top 20 PCs, and recruitment centers (as categorical variables) in all association analyses. We ran additional sensitivity analyses accounting for 40 PCs to assess potential residual population stratification and found the exome-wide burden results remained consistent (Table S2). The effect size (Beta), 95% confidence interval and p-values were calculated for each burden association. The significance level of the burden association was determined by Bonferroni correction for the number of association tests within the defined set of analysis and is provided in the respective tables.

#### Exome-wide burden

To characterize the effects of exome-wide rare coding variant burden on cognitive function, we calculated the cumulative minor allele counts of rare coding variants (MAF<1.0×10^-5^) for each variant functional class defined by VEP ^29^, LOFTEE ^33^, MPC ^32^ and pLI scores ^33^. We defined the following variant classes: protein truncating variants (PTV; high confidence loss-of-function (LoF) variants identified by VEP with LOFTEE plugin); missense variants (identified by VEP) classified by deleteriousness (MPC score) into tier 1 for MPC>3, tier 2 for 3≥MPC>2 and tier 3 for other missense variants not in the previous two tiers; synonymous variants (identified by VEP). We further classified variants by LoF intolerance of the gene (pLI≥0.9 or pLI<0.9) in which the variant resides. For each UKB sample, the exome-wide rare coding variant burden for each variant class was calculated. We performed exome-wide burden association in EUR, SAS and AFR samples in UKB. However, the sample size of SAS (N=7,086) and AFR (N=6,330) unrelated samples is much smaller compared with EUR unrelated samples (N=321,843) and we did not replicate the exome-wide burden association signals we identified in EUR using the SAS and AFR samples. A down-sampled exome-wide burden analysis (N=7,000) in EUR also did not replicate the finding in full EUR sample (data not shown).

#### Gene set burden - ASD, DD, DDG2P exome genes

We examined the impact of rare coding variants in genes identified in autism spectrum disorder (ASD; N=102) ^11^ and developmental disorder (DD; N=285) ^13^ exome studies, as well as in DD genes listed in the Development Disorder Genotype - Phenotype Database (DDG2P; https://www.deciphergenomics.org/ddd/ddgenes). DDG2P provides a curated list of genes reported to be associated with developmental disorders and curated by clinicians as part of the Deciphering Developmental Disorders (DDD) study to facilitate clinical reporting of likely causal variants. We included 2,020 confirmed and probable DD genes from DDG2P into our analysis. ASD, DD and DDG2P gene set burdens were calculated, and burden association analyses were conducted following the analysis procedure described above in unrelated UKB EUR samples (N sample=321,843).

#### Gene set burden - GWAS genes

Following the same procedure as above, we performed gene set-based burden analysis in genes identified in GWAS for educational attainment (N gene=1,140) ^5^, cognitive function (N gene=807) ^48^, schizophrenia (N gene=3,542) ^76^, bipolar disorder (N gene=218) ^77^ and depression (N gene=269) ^78^. The respective GWAS gene lists were taken either directly from publications or from reprocessing the publicly available GWAS summary statistics with FUMA for positional gene mapping ^79^. We also calculated rare coding variant burden for a set of randomly selected genes not linked to cognitive function or psychiatric disease (N gene=1,082), which includes 5% of all genes excluding cognitive function genes identified in this study, ASD and DD exome studies ^12, 13^ and cognitive function, educational attainment, schizophrenia, bipolar disorder and depression GWAS ^5, 48, 76–78^. We followed the same procedure in exome-wide burden analysis to perform the gene set burden association analysis in unrelated UKB EUR samples.

#### Gene set burden - MSigDB curated gene sets and pathways

To identify pathways and gene sets associated with cognitive function, we calculated PTV burden for 13,011 gene sets identified in the Molecular Signatures Database (MSigDB v7.2; accessed on 11/26/2020) and performed self-contained pathway association analysis^80^. We included all C2 canonical pathways (BioCarta N gene=292; KEGG N gene=186; Pathway Interaction Database [PID] N gene=196; Reactome N gene=1,547; WikiPathways N gene=587) and C5 gene ontology (GO) pathways (Biological Process N gene=7,531; Cellular Component N gene=996; Molecular Function N gene=1,676). Again, we followed the same procedure as above to examine the association between pathway PTV burden and cognitive function phenotypes. The significance level was determined by Bonferroni correction for the number of pathways and gene sets tested for each phenotype (0.05/13011 = 3.84×10^-6^). Note that we performed self-contained pathway analysis where the null hypothesis is that there is no association between the gene set PTV burden and the cognitive function phenotype (as opposed to competitive pathway analysis) ^80^.

#### Gene set burden - Genes with brain specific and non-specific expression

To examine a potential enrichment of rare coding variant burden association among genes with brain specific expression in cognitive function, we calculated rare coding variant burdens for three gene sets defined by gene expression specificity in the Human Brain Atlas ^81^: genes with elevated expression in brain (N gene=2,587); genes with elevated expression in other tissues, but also expressed in brain (N gene=5,298); and genes with expression that has low tissue specificity (N gene=8,385). The burden association analysis was done in unrelated European UKB samples with the same procedure as above.

#### Exome-wide gene-based PTV burden test

To identify genes associated with adult cognitive function, we calculated the rare PTV burden for each gene and performed burden association analyses. We used two-step whole genome regression implemented in Regenie for association testing ^35^. Regenie accounts for population stratification and sample relatedness, which allowed us to leverage a larger sample size by including related samples. Regenie first fits a stacked block ridge regression to obtain leave-one-chromosome-out (LOCO) genetic prediction of the phenotype of interest and in the second step, the association test is carried out by fitting regression models conditioning on the LOCO predictions derived in the first step.

For the Regenie step one regression, we first performed sample QC (as described in the earlier section) and QC on UKB genotype data by excluding variants with genotyping call rate less than 90%, Hardy-Weinberg equilibrium test p-value smaller than 10^-15^ and MAF smaller than 1%. This retained 565,124 genotyped variants for the step one regression. We fitted Regenie first step regression for IRNT EDU, RT and VNR separately, adjusting for sex, age, age^2^, sex by age interaction, sex by age^2^ interaction, top 20 PCs, and recruitment centers in UKB European samples (identified with in-house population assignment described in the previous section, maximum N with exome sequencing=397,434) with 10-fold cross validation. We used Regenie version 1.0.6.7 for the step one regression.

For the Regenie step two association test, we implemented an in-house R pipeline for rare PTV burden association tests conditioned on the first step LOCO prediction, following the linear regression model for association testing described in the Regenie publication ^35^. We treated the LOCO prediction as an offset in the linear regression model where IRNT EDU, RT and VNR were regressed on gene-based rare PTV burden, adjusting for the same covariates used in step one regression. We excluded genes with less than 10 PTV carriers from the gene-based PTV burden analysis, which leads to variable number of tests performed for each phenotype (especially for VNR that has much smaller sample size). The sample sizes and number of genes tested for each cognitive function phenotype were as follows: N sample=393,758 and test N=15,782 for EDU; N sample=394,600 and test N=15,798 for RT; N sample=159,026 and test N=11,905 for VNR. The Bonferroni correction for multiple testing was based on the actual number of tests performed per phenotype and across three phenotypes. The significance levels for the gene-based rare PTV burden association tests were Bonferroni corrected for the number of tests for each phenotype separately, which are 0.05/15,782=3.17×10^-6^ for EDU, 0.05/15,798=3.16×10^-6^ for RT and 0.05/11,905=4.20×10^-6^ for VNR. We note that 7 out of the 8 cognitive function genes (i.e., all except *BCAS3*) identified in our PTV burden association analysis were also exome-wide significant after Bonferroni correction across all tests (*P*<0.05/43,485=1.15×10^-6^). We identified 5 additional genes with FDR Q-value less than 0.05 for EDU and VNR (Fig. 1 and Table S5).

We note that some of the findings reported in our study have also been observed in a recent cross-phenotype exome study by Backman et al. ^21^, where gene-based burden analyses for RT were conducted using different “gene burden masks” (annotations for grouping variants to calculate burden) and MAF cut-offs than our analysis. For verbal-numerical reasoning (originally termed “fluid intelligence” by UK Biobank), Backman et al. formatted the participants’ responses to each verbal-numerical reasoning question into binary traits using one of the multiple choices against other choices and examined each question separately in their gene-based burden analyses. This is not consistent with how the UKB “fluid intelligence” data were analyzed in previous cognitive function GWAS ^2, 3, 48^. The total number of correctly answered questions as we analyzed in this study consistent with the phenotype definition used in previous cognitive function GWAS and is considered a standard measure of general cognitive function ^2, 3, 48^.

#### The SUPER-Finland study

To replicate our findings, we tested association between the genes identified in our gene-based PTV burden analysis (8 genes with Bonferroni corrected significance and 13 with FDR significance) and cognitive traits in the SUPER-Finland study. The SUPER-Finland study is a cohort of 9,125 psychotic patients in Finland. Subjects with a diagnosis of a schizophrenia spectrum psychotic disorder (ICD-10 codes F20, F22-29), bipolar I disorder (F31) or major depressive disorder with psychotic features (F32.3 and F33.3) were recruited from in-and outpatient psychiatric, general care and housing units and by advertisements in local newspapers. DNA samples were genotyped with GWAS arrays, exome sequenced and linked to a wide range of phenotypic information ascertained through a structured interview, questionnaires, and cognitive testing. After receiving written and verbal information on the study and biobank research, all participants gave written informed consent for participating in the study. The sample collection was conducted between 2016 and 2018 and has been funded by the Stanley Center for Psychiatric Research at the Broad Institute of MIT and Harvard, Boston, USA, and forms one arm of the Stanley Global Neuropsychiatric Genomics Initiative.

#### Whole-exome sequencing data generation and quality control

Blood samples were collected by venipuncture for DNA extraction (2x Vacutainer EDTA K2 5/4 ml, BD, serum (Vacutainer STII 10/8 ml gel, BD) and plasma (Vacutainer EDTA K2 10/10 ml, BD) analyses. In cases where venipuncture was not possible, a saliva sample (DNA OG-500, Oragene) was collected for DNA extraction. DNA extraction from EDTA-blood tubes was performed using PerkinElmer Janus chemagic 360i Pro Workstation with the CMG-1074 kit. Saliva samples (n= 509) were incubated in +50°C overnight before DNA extraction. Saliva samples were processed using Chemagen Chemagic MSM I robot with CMG-1035-1 kit. DNA was eluted in 400 µl 10 mM Tris-EDTA elution buffer (PerkinElmer) and DNA-concentration measured with Trinean DropSense spectrophotometer. Samples were aliquoted with Tecan Genesis/Tecan Freedom Evo and shipped to the Broad institute of MIT and Harvard, Boston, USA on dry ice for genetic analyses.

Exome sequencing was performed at the Broad Institute. The sequencing process included sample prep (Illumina Nextera, IIlumina TruSeq and Kapa Hyperprep), hybrid capture (Illumina Rapid Capture Enrichment (Nextera) - 37Mb target and Twist Custom Capture - 37Mb target) and sequencing (Illumina HiSeq4000, Illumina HiSeqX, Illumina NovaSeq6000 - 150bp paired reads). Sequencing was performed at a median depth of 85% targeted bases at > 20X. Sequencing reads were mapped by BWA-MEM to the hg38 reference using a “functional equivalence” pipeline. The mapped reads were then marked for duplicates, and base quality scores were recalibrated. They were then converted to CRAMs using Picard and GATK. The CRAMs were then further compressed using ref-blocking to generate gVCFs. These CRAMs and gVCFs were then used as inputs for joint calling. To perform joint calling, the single-sample gVCFs were hierarchically merged (separately for samples using Nextera and Twist exome capture).

Quality control (QC) analyses were conducted in Hail 0.2 and the full details are described in the SCHEMA manuscript. In brief, we annotated variants as frameshift, inframe deletion, inframe insertion, stop lost, stop gained, start lost, splice acceptor, splice donor, splice region, missense, or synonymous using the Ensembl Variant Effect Predictor tool. At the genotype level, we first split multiallelic sites and retained individual calls if they had a genotype quality (GQ) ≥ 20, allelic balance (AB) < 0.1 in homozygous calls, allelic balance (AB) ≥ 0.25 in heterozygous calls and depth (DP) ≥ 10. After applying genotype filters, we excluded variants with call rates < 0.9 or if they resided within low-complexity regions (LCR). We excluded samples that were 4 median absolute deviations from the mean in any of the following metrics: call rate (callRate), number of heterozygous calls (nHet), number of homozygous alternate calls (nHomVar), number of non-reference calls (nNonRef), number of deletions (nDeletion), number of insertions (nInsertion), number of singleton calls (nSingleton), number of SNPs (nSNPs), heterozygous-homozygous call ratio (rHetHomVar), transition-transversion ratio (rTiTv) and insertion-deletion (rInsertionDeletion). Using the Hail PC-relate function, we pruned clusters of related individuals to ensure that no two samples were second-degree or closer in relations. Individuals with a Finnish predicted ancestry (P > 0.7) using a Random Forest model based on PCA using 1000 Genomes as a basis were retained.

#### Cognitive function phenotypes for replication analysis

The SUPER-Finland study protocol included a questionnaire, a structured interview by a research nurse, physical measurements, and blood/saliva sampling. The questionnaire and interview included questions on educational attainment (http://www.julkari.fi/handle/10024/78534), academic performance and learning difficulties at school, which were derived from the Finnish Health 2000 and 2011 general population surveys. We identified PTV carriers in the cognitive genes in the primary analysis and performed association tests between cognitive gene PTV burden and cognitive phenotypes. Association tests were done with either linear or logistic regression. We regressed the phenotypes on PTV status and corrected for 10 principal components, imputed sex, sequence assay and total number of coding variants in the genome. We focused on the following phenotypes: we define a developmental disorders/intellectual disability (DD/ID) case as someone with a diagnosis of learning difficulty and intellectual disability based on diagnostic codes in HILMO (THL). In the interview data, we asked “How did you fare in studies compared to your schoolmates?” with a response encoding of ‘Below average’, ‘Moderately’, ‘Better than average’. Level of education completed was a trinarized measure based on definitions in the Health 2000 survey, and is encoded as ‘low’, ‘middle’, ‘high’ and correlates with only having completed primary, secondary, and tertiary education.

#### The Northern Finland Intellectual Disability (NFID) study

To replicate our gene findings, we performed PTV burden association analysis for the genes identified in our UKB PTV burden analysis in 1,097 intellectual disability cases from Northern Finland Intellectual Disability (NFID) study and 11,774 controls from the FINRISK 1992-2012 and Health 2000-2011 studies ^36^. The details of the study sample recruitment and phenotyping, exome sequencing data generation and quality control and ethical permissions were described in Kurki et al. 2019 ^36^. Specific to the replication analysis in the current study, we tested association between gene set PTV burden (8 or 13 genes identified in UKB gene-based PTV burden analysis) and risk for DD/ID in a logistic regression model, adjusted for sex and top 10 PCs.

#### The Mass General Brigham Biobank

To replicate our gene discovery findings, we tested association between the genes identified in our gene-based PTV burden analysis (8 genes with Bonferroni corrected significance and 13 with FDR significance) and educational attainment in participants of the Massachusetts General Brigham Biobank (MGBB) with whole-exome sequencing data. The MGBB is a hospital-based biobank aiming at collecting blood samples, lifestyle and family history survey data, as well as electronic health record linkage from consented participants ^82^. The release used for this study (as of November 2021) includes 24,787 samples that were whole-exome sequenced and genome-wide genotyped in two batches. All MGBB patients gave informed consent for general biobank research.

#### Genotype and sample quality control

We conducted QC of genome-wide genotypes for 24,787 samples following a QC pipeline (https://github.com/Annefeng/PBK-QC-pipeline) by using PLINK1.9, R, and python scripts. The following filters were used in sequence: variant call rate>0.95; sample call rate>0.98; second round variant filter with call rate>0.98. Variant-level missing rate was computed in each batch and variants with missing rate difference > 0.75% were filtered out. After merging two genotyping batches, we further removed duplicated variants, monomorphic variants and variants not confidently mapped to any chromosomes.

To identify MGBB samples of European ancestry, we leveraged 1000 Genomes (1KG) Project phase 3 samples as population reference. To do so, we first combined MGBB genotypes with 1KG genotypes (N sample=2,504) ^74^. We only retained overlapping variants with MAF>0.05 and call rate>0.98 and filtered out multi-allelic and strand ambiguous variants. We LD-pruned variants at R^2^ = 0.1 with window size 200 kb to obtain independent variants for principal components analysis (PCA), while excluding variants in long-range LD regions (chr6:25-35Mb and chr8:7-13Mb). With 1KG super population labels (European [EUR], East Asian [EAS], African [AFR], American [AMR] and South Asian [SAS]), we used top 6 PCs to train a random forest model and assigned MGBB samples into five populations (prediction probability>0.8). We identified 17,287 (69.7%) EUR samples for the subsequent analysis.

We further QCed MGBB EUR samples by filtering out 513 samples, including samples whose reported sex was different from genetically imputed sex (F-statistics<0.2 imputed as female; F-statistics>0.8 as male), samples with outlying heterozygosity rate (>5 standard deviation from the mean) and one of each pair of related samples (pi-hat>0.2). After removing variants showing significant batch effects (*P*<1.0×10^-4^), we performed PCA of QC-ed EUR samples and removed 73 outlier samples (6 standard deviations away from the mean in top 10 PCs). A total of 16,701 samples of European ancestry were retained as the final analytical sample. In-sample PCs were used to control for population stratification in the replication analysis.

#### Whole-exome sequencing data generation and quality control

Whole-exome sequencing was done in 26,421 MGBB samples using Illumina NovaSeq with a custom exome panel (TWIST Biosciences). The sequencing coverage was 20X for more than 85% of exonic target. Variants were joint-called by Genome Analysis ToolKit (GATK) GVCF workflow with HaplotypeCaller in gVCF mode. WES data quality control was done with Hail v0.2 (https://github.com/hail-is/hail). We first split multi-allelic variants into bi-allelic and retained high-quality variants with variant level genotype quality (GQ)>20, call rate>0.9, allele count>0, 200>mean depth (DP)>10, allele balance (AB)>0.9. Then, variants were separated into SNPs and indels for hard filtering. For SNPs, we kept SNPs with QualByDepth (QD)≥2, FisherStrand (FS)≤60, StrandOddsRatio (SOR)≤3, RMSMappingQuality (MQ)≥40, MappingQualityRankSumTest (MQRankSum)≥-12.5 and ReadPosRankSumTest (ReadPosRankSum)≥-8. For indels, we kept variants with QD≥2, ReadPosRankSum≥-20, FS≤200 and SOR≤10. We retained 10,588,646 high-quality variants after QC. With high-quality variants, we then performed sample-level QC by keeping samples with number of singleton (n.singleton)<500, sample-level genotype quality (GQ)>40 and sample call rate>0.9.

#### Statistical analysis

We performed replication analysis on MGBB samples of European ancestry with genotype, whole-exome sequencing and educational attainment data using R. The total analytic sample size is 8,389. We first extracted variants in the 8 genes identified in UKB gene-based PTV burden analysis from the MGBB exome data. We annotated the coding variants with Variant Effect Predictor (VEP) v96 ^29^ and Loss-Of-Function Transcript Effect Estimator (LOFTEE) ^30^. We identified 28 PTVs in the 8 genes and 36 PTVs in 13 genes with MAF ranging from 1.89×10^-5^ to 7.57×10^-5^. We calculated PTV burden across 8 or 13 cognitive genes in MGBB European samples and performed association tests between cognitive gene PTV burden and educational attainment. Educational attainment in MGBB was self-reported and converted from categories of educational levels to years-of-education following the sample rules used in processing UKB educational attainment data. We used linear regression for association testing, adjusted for sex, age, age^2^, sex by age interaction, sex by age^2^ interaction and top 20 PCs.

#### Phenome-wide association analysis

To explore the cognitive function genes identified for potential pleiotropic effects, we performed a PTV burden phenome-wide association analysis across 3,150 semi-automatically derived UKB phenotypes. The binary phenotypes include ICD10 codes from in-patient records (ICD-10 congenital malformations; deformations and chromosomal abnormalities; ICD-10 diseases of the circulatory system; ICD-10 diseases of the digestive system; ICD-10 diseases of the eye and adnexa; ICD-10 diseases of the genitourinary system; ICD-10 diseases of the musculoskeletal system and connective tissue; ICD-10 diseases of the nervous system; ICD-10 diseases of the respiratory system; ICD-10 diseases of the skin and subcutaneous tissue; ICD-10 endocrine, nutritional and metabolic diseases; ICD-10 mental, behavioral and neurodevelopmental disorders; ICD-10 neoplasms; ICD-10 pregnancy, childbirth and the puerperium; ICD-10 symptoms, signs and abnormal clinical and laboratory findings, not elsewhere classified) and death records (ICD-10 cause of death), self-reported illness (cancer and non-cancer), self-reported medication, surgery/operation codes and family history (fathers’ illnesses, mothers’ illnesses and siblings’ illnesses were combined into a single phenotype for each of the 12 family history illnesses ascertained for in UK Biobank questionnaires). The quantitative phenotypes include biomarkers such as blood cell count, blood biochemistry, infectious disease antigen assays and physical measurements. A list of all phenotypes with phenotype categories, UKB field numbers and phenotype full names can be found in Table S7.

We restricted the phenome-wide association analysis to 321,843 unrelated UKB participants of European ancestry. We excluded binary phenotypes with fewer than 100 cases in our analysis. PTV burden test for binary phenotypes was performed using logistic regression in all individuals, controlling for sex, age, age^2^, sex by age interaction, sex by age^2^ interaction, top 20 PCs, and assessment centers. For binary phenotypes with PTV burden association p-value < 0.01, we repeated the analysis using the Firth logistic regression to account for situations where the logistic regression outputs may be biased due to separation^83^. For quantitative phenotypes, we excluded phenotypes with fewer than 100 observations. For each quantitative phenotype, individuals with outlier phenotype values (> 5 SD from the mean) were excluded. PTV burden test for quantitative traits was performed using linear regression on inverse rank-based normal transformed (IRNT) phenotypes in all individuals, controlling for sex, age, age^2^, sex by age, sex by age^2^, top 20 PCs and assessment centers. We defined a Bonferroni-corrected phenome-wide significance threshold by the number of tests per gene of 1.59 x 10^-5^ (0.05/3,150).

#### Characterization of cognitive phenotypes in KDM5B and CACNA1A PTV carriers

*KDM5B* is an established Mendelian disease gene, with homozygous or compound heterozygous mutations causing autosomal recessive intellectual disability (OMIM #618109) ^45, 46^. Similarly, *CACNA1A* is also an established disease gene with heterozygous mutations causing developmental and epileptic encephalopathy (OMIM # 617106) ^42, 43^, type 2 episodic ataxia (OMIM # 108500) ^41^, or spinocerebellar ataxia (OMIM # 183086) ^39, 40^. To better understand the relationship between PTVs in *KDM5B* and *CACNA1A* and cognitive function phenotypes, we first processed the EDU and VNR in the UKB European samples by residualizing EDU and VNR with sex, age, age^2^, sex by age interaction, sex by age^2^ interaction, top 20 PCs and recruitment centers and then standardized the residuals by inverse rank-based normal transformation. Then, we plotted the standardized, residualized EDU and VNR for each PTV carrier against the genomic position of the PTV to characterize the phenotype distribution for *KDM5B* and *CACNA1A* PTV carriers (Fig. 3 and Fig. S6). We further compared the standardized, residualized EDU and VNR between 3 groups of PTV carriers for *KDM5B* and *CACNA1A*: 1) PTV carriers who do not have any in-patient ICD diagnostic code records for neurological, psychiatric, or neurodegenerative disorders or carry Clinvar pathogenic/likely pathogenic variants; 2) PTV carriers with in-patient ICD diagnostic code records for neurological, psychiatric, or neurodegenerative disorders; and 3) PTV carriers of ClinVar pathogenic/likely pathogenic variants (Table S10 and S11).

#### Temporal expression of cognitive function genes

We obtained temporal RNA-seq expression data from BrainSpan ^47^, an atlas of the developmental human brain. This data was generated from 42 individual donors, across 26 brain regions and in 31 developmental ages, with 524 samples in total. Gene expression was originally processed as reads per kilobase per million (RPKM). We first removed genes that were not expressed (RPKM<1) in more than 10% of the total samples, resulting in 11,744 genes with expression information available. Then, RPKM was transformed to log_2_ (RPKM+10^-8^) (adding 10^-8^ to avoid possible numerical error in logarithm transformation). Thirty-one developmental ages were grouped into 8 developmental stages, early prenatal (8-12 pcw), early mid-prenatal (13-18 pcw), late mid-prenatal (19-24 pcw), late prenatal (25-38 pcw), infancy (0-18 months), childhood (19 months - 11 years), adolescence (12-19 years) and adulthood (20-60+ years). Temporal expression of the cognitive function genes across development stages was fitted loess regression. We compared prenatal and postnatal expression of the cognitive function genes across the QCed dataset with a two-sided two-sample student’s *t*-test. We also performed one-way ANOVA to test if the means of different developmental stages were significantly different. A post-hoc Tukey multiple pairwise-comparison between the means of stages was conducted if one-way ANOVA showed significant results.

#### Kdm5b mouse model and RNA-seq

##### RNA extraction, sequencing, and data processing

Mouse tissues were homogenized in buffer RLT plus (Qiagen) with β-mercaptoethanol (Sigma, M3148; 10µl/ml) using Qiagen TissueLyser LT, with sterile steel beads and operated at 50Hz for 2 minutes. Samples were passed over gDNA eliminator columns. Then total RNA was extracted on RNeasy Plus columns as per manufacturer’s protocol (Qiagen), immediately snap frozen on dry ice and stored at −80C. An aliquot of each sample was quantified using 2100 Bioanalyzer (Agilent Technologies). RNA sequencing libraries were prepared using established protocols: library construction (poly(A) pulldown, fragmentation, 1st and 2nd strand synthesis, end prep and ligation) was performed using the NEB Ultra II RNA custom kit (New England Biolabs) on an Agilent Bravo automated system. Indexed multiplexed sequencing was performed on an Illumina HiSeq 4000 system, using 75 bp paired-end sequencing reads. The sequencing data were de-multiplexed into separate CRAM files for each library in a lane. Adapters that had been hard-clipped prior to alignment were reinserted as soft-clipped post alignment, and duplicated fragments were marked in the CRAM files. The data pre-processing, including sequence QC and STAR alignments were made with a Nextflow pipeline, which is publicly available at https://github.com/wtsi-hgi/nextflow-pipelines/blob/rna_seq_mouse/pipelines/rna_seq.nf, including the specific aligner parameters. We assessed the sequencing data quality using FastQC v0.11.8. Reads were aligned to the GRCm38 mouse reference genome (Mus_musculus.GRCm38.dna.primary_assembly.fa, Ensembl GTF annotation v99). We used STAR version 2.7.3a^84^ with the --twopassMode Basic parameter. The STAR index was built against Mus_musculus GRCm38 v99 Ensembl GTF using the option -sjdbOverhang 75. We then used featureCounts version 2.0.0 ^85^ to obtain a count matrix. Genes with less than 5 counts in more than 33% of samples were filtered out. The counts were normalized using DESEQ2’s median of ratios method ^86^. Differential gene expression and log2 fold changes were obtained using the DESEQ2 package ^86^ with SVA correction ^87^. The default DESEQ2 adjusted p-value threshold of 0.10 was used to identify significant differences between wildtype and mutant samples. The number of differentially expressed genes (DEG) in each tissue was considered as the union of DEG in both *Kdm5b^+/-^* and *Kdm5b^-/-^* animals.

For the identification of functionally enriched terms in the differentially expressed genes, Gene ontology (GO) enrichment analysis was performed using the gprofiler R package (gost function, ordered_query = FALSE). A threshold of 5% FDR and an enrichment significance threshold of P<0.05 (correction_method = “fdr” for multiple testing) was used. In all analyses, the background consisted of only the genes considered expressed in the tissue studied (genes that passed the minimum count filtering that had adjusted p-value with a numerical value, different to NA). GO terms with more than 1,000 genes were excluded from the analysis. The European Nucleotide Archive accession numbers for the RNA-seq sequences reported in this paper are as listed in Table S15.

#### Animals

##### Generation of *Kdm5b* loss of function mice

A mouse *Kdm5b* loss of function allele (MGI:6153378) was generated previously^45^ by CRISPR/CAS9 mediated deletion of coding exon 7 (ENSMUSE00001331577), leading to a premature translational termination due to a downstream frameshift. Breeding of testing cohorts was performed on a C57BL/6NJ background. Breeding, housing of mice and all experimental procedures with mice were assessed by the Animal Welfare and Ethical Review Body of the Wellcome Sanger Institute and conducted under the regulation of UK Home Office license (P6320B89B), and in accordance with institutional guidelines. Mice were housed in mixed genotype cages (2-5 mice) with food and water ad-libitum, under controlled temperature and humidity and a 12h light cycle (light on at 7:30am) at the Research Support Facility of the Wellcome Sanger Institute.

##### Behavioral testing

We applied a battery of behavioral tests as they are commonly applied to study mice for signs of perturbed neurodevelopment. Specifically, we assessed a cohort of 25 wildtype, 34 heterozygous and 15 homozygous *Kdm5b* mutant male mice at 10 weeks of age. Behavioral tests were carried out between 9 am – 5 pm, after 1 hr of habituation to the testing room. Experimenters were blind to genotype, and mouse movements were recorded with an overhead infrared video camera for later tracking by automated video tracking (Ethovision XT 11.5, Noldus Information Technology).

###### Light-dark box

This test was adapted from Gapp et al. ^88^. Mice were housed in pairs or trios for at least 10 minutes before performing a grip strength test (BIO-GS3, Bioseb) and subsequently introducing them individually into the light-dark box, a plastic box (40 × 42 × 26 cm) divided in two compartments. One is smaller, closed, and dark (1/3 of the total surface area) and connected through a door (5 cm) to a larger, brightly lit (370 lux with an overhead lamp) compartment (2/3 of the box). Each mouse was placed in the dark compartment, the door was then opened, and the animal was left to explore for 10 min. The time spent in the light compartment was assessed and the difference between genotype groups was calculated as z-scores.

###### Barnes Maze probe trial

This assay is a test of visuo-spatial learning and memory on a circular maze (120 cm diameter table) with 20 holes around the perimeter ^89^. One of the holes leads to a small dark box (Target) where the mice can escape from the brightly lit maze. Mice were trained for three days, 10 trials (4 min maximum each), to find the target location. On the probe trial, 72 hr after the last training day, the escape box was removed. Each mouse was given 4 minutes to explore the maze. The mouse’s movements were tracked, and the amount of time spent around each of the holes during the first minute of the test was analyzed. Analysis results are expressed as z-scores of homozygote or heterozygote relative to wildtype mice.

###### Novel Object Recognition

This assay was conducted as part of an Object Displacement - Novel Object Recognition test on a square arena (37 cm side). Mice were first habituated to the arena for 20 min. The following day mice were tested during two 10min trials, with an inter-trial-interval of 1 hr. During these trials, mice were left to explore two identical objects (either small glass bijou bottles or similarly sized halogen light bulbs). 24 hr later, on day 3, mice were given the choice to explore two different objects, a familiar one (to which they had been exposed the previous day) and a novel one. The movement of each mouse was tracked and the amount of time their nose was in proximity to the objects was recorded and used as investigation time. The preference for investigating the novel object was calculated as a ratio, Preference Novel= T*novel/*(T*novel* + T*familiar*), where T*novel* and T*familiar* are the amount of time spent investigating the novel and familiar objects. Mice prefer to investigate novel objects over familiar ones, so deviation from this bias is interpreted as reduced recognition memory sensitivity or discriminatory ability ^90^.

###### X-rays

Fifteen *Kdm5b^+/+^*, twelve *Kdm5b^+/-^*and nine *Kdm5b^-/-^*mice were anesthetized with ketamine/xylazine (100mg/10mg per kg of body weight) and then placed in an MX-20 X-ray machine (Faxitron X-Ray LLC). Whole body radiographs were taken in dorso-ventral and lateral orientations. Images were then analyzed, and morphological abnormalities assessed using Sante DICOM Viewer v7.2.1 (Santesoft LTD). To sufficiently power the analysis of transitional vertebrae in heterozygous animals, we analyzed a larger number of animals (*Kdm5b^+/+^:* 46*, Kdm5b^+/-^: 40, Kdm5b^-/-^*:21).

#### Statistical analysis

All statistical analyses of mouse data were performed using R. Data was first transformed to achieve normality, using Box Cox transformation (MASS package) for behavior data (lambda limit = [-2,2]) or quantile normalization (qnorm function) for x-ray data. Testing for genotype effect was performed using a double generalized linear model, dglm (dglm package). The type of object used for Novel Object Recognition had a small (6%) and significant (*P*=0.036) effect, therefore was used as a covariate for Box Cox transformation and dglm. For visualization purposes, residual values were calculated from the linear model and z-scores relative to wildtypes were calculated.

### Overlapping rare and common variant association signals

To compare and contrast the genetic loci identified through common variant association tests in GWAS to genes identified in our rare PTV burden analysis, we cross-checked all independent genome-wide significant variants in the most recent, largest educational attainment GWAS ^5^ and cognitive function GWAS ^48^ with the 13 cognitive function-associated genes identified in our exome analysis. For the educational attainment GWAS, we assessed any independent genome-wide significant variants listed in Lee et al. 2018 ^5^ Supplementary Table 2 where the PTV burden identified cognitive function genes are located in nearby regions. We identified one SNP, rs10798888 (chr1:31733498 [GRCh38]), with a minor allele frequency of 0.1725 and an association p-value of 5.15×10^-14^ with educational attainment in the region where *ADGRB2* PTV burden showed an association with EDU (*P*=8.55×10^-12^). We then extract the surrounding region of SNP rs10798888 from the full summary statistics of educational attainment GWAS (excluding 23andMe data) obtained from the Social Science Genetic Association Consortium (SSGAC; https://thessgac.com/), generated regional plots (https://my.locuszoom.org/) ^91^ of the GWAS association results and compared these with the cognitive function phenotypes (EDU and VNR) among PTV carriers in UKB EUR samples (Fig. S11). The variants in GWAS regional plots were further annotated for previous GWAS associations registered in the GWAS catalog with LocusZoom’s automated annotation feature.

For cognitive function GWAS, we processed the GWAS summary statistics from Lam et al. 2021 ^48^ with a GWAS summary statistics quality control pipeline ^92^ and used FUMA ^79^ to identify independent genome-wide significant loci for cognitive function from the GWAS. We identified one genome-wide significant locus on chromosome 22 with the top independent genome-wide significant SNP rs5751191 (chr22:41974987 [GRCh38], association *P*=2.02×10^-12^) that overlapped with *NDUFA6* which showed FDR significant association with EDU (*P*=6.98×10^-6^, FDR *Q*=0.016) in our PTV burden association analysis in UKB EUR samples. We extracted variants in the region that covered the variants in LD with the top SNP rs5751191 (R^2^>0.6) to generate a regional plot and identified genes in the region to extract the corresponding PTV-burden association test p-values and number of PTV carriers in UKB EUR samples.

### Contributions of common variants and rare coding variants on EDU and VNR

We examined the relative contribution of common variants and rare damaging coding variants on cognitive function. To do so, we first calculated polygenic risk scores (PRS) to capture the impact of genome-wide common variants on cognitive function, using imputed genome-wide genotypes and variant weights derived using PRS-CS ^49^ based on a cognitive function GWAS meta-analysis ^48^. The cognitive function GWAS meta-analysis included only European samples and was done using the latest cognitive genomics consortium (COGENT) data freeze excluding samples from UKB ^48^. We applied polygenic risk scores-continuous shrinkage (PRS-CS) ^49^ to derive the variant weights with a pre-computed LD reference panel based on 1000 Genomes Project phase 3 European superpopulation samples. The PRS-CS global shrinkage parameter phi was set to be 0.01 because cognitive function is a known highly polygenic trait. Using PRS-CS derived variant weights and QCed imputed genotypes, we calculated PRS as a weighted sum of counted alleles across the genome using PLINK 2.00. Second, following the exome-wide burden analysis, we identified rare damaging coding variant carriers as carriers of rare PTV and/or damaging missense variants with MPC>2 in LoF intolerant genes (pLI>0.9) across the exome.

To demonstrate the relative impact of PRS and rare damaging coding variant carrier status, we plotted standardized, residualized EDU and VNR against PRS, stratified by rare damaging coding variant carrier status in unrelated UKB European samples. The phenotypes were residualized by sex, age, age^2^, sex by age interaction, sex by age^2^ interaction, top 20 PCs, and recruitment centers and then inverse rank-based normal transformed. The samples were grouped by PRS in 2% quantiles. The median of standardized, residualized EDU and VNR was calculated and plotted for each PRS group. We further assess the prediction performance of cognitive function PRS and rare damaging coding variant carrier status for EDU and VNR. We fitted linear regression models by regressing IRNT EDU and VNR on PRS and rare damaging coding variant carrier status jointly, adjusted for covariates, in unrelated UKB European samples. The regression coefficients, association p-value and partial R^2^ were estimated ^93^. We further examined the interaction between PRS and rare damaging coding variant carrier status by adding an interaction term to the linear regression model and tested for significant interaction effects. We also modeled PRS as a continuous variable and a binary variable by dividing samples in the top 10% PRS group vs the rest 90% PRS group.

## Data Availability

All data produced in the present study are available upon reasonable request to the authors.

## Data availability

pLI score is available at https://storage.googleapis.com/gcp-public-data--gnomad/release/2.1.1/constraint/gnomad.v2.1.1.lof_metrics.by_gene.txt.bgz MPC score is available at ftp://ftp.broadinstitute.org/pub/ExAC_release/release1/regional_missense_constraint/ Brainspan RNA sequencing data is available at https://www.brainspan.org/static/download.html Human protein atlas data are available at https://www.proteinatlas.org/humanproteome/brain/human+brain The Development Disorder Genotype - Phenotype Database (DDG2P) gene list is available at https://www.deciphergenomics.org/ddd/ddgenes Social Science Genetic Association Consortium (SSGAC) educational attainment GWAS summary statistics are available at https://www.thessgac.org/ (registration required)

## Code availability

PRS-CS: https://github.com/getian107/PRScs

PLINK2.00: https://www.cog-genomics.org/plink/2.0

VEP: https://useast.ensembl.org/info/docs/tools/vep/index.html

LOFTEE: https://github.com/konradjk/loftee

Regenie: https://rgcgithub.github.io/regenie/

Hail 0.2: https://github.com/hail-is/hail

LocusZoom: https://my.locuszoom.org/

## Acknowledgements

We thank all the participants and researchers of the UK Biobank, Mass General Brigham Biobank, the SUPER-Finland study, and the Northern Finland Intellectual Disability (NFID) study. The use of UK Biobank data (whole-exome sequencing, genome-wide genotyping, and phenotypic data) in the current study is approved under application #26041. We thank Mass General Brigham Biobank for providing genomic and health information data. The healthy control samples/data used in the NFID study analysis were obtained from THL Biobank. We thank all study participants for their generous participation at THL Biobank and FINRISK 1992-2012 and Health2000-2011 studies. We thank the staff from the Wellcome Sanger Institute’s Research Support Facility for mouse husbandry and support, Sanger Institute Sample Management and DNA Pipelines for providing the RNA sequencing and Vivek Iyer and Guillaume G Noell (Human Genetics Informatics, Wellcome Sanger Institute) for Bioinformatic support.

## Author contributions

C.-Y.C. and H.R. conceived and supervised the study. C.-Y.C., R.T., T.G., M.L., T.S., L.U., J.Z.L. performed the analyses. G.S.A., M.S., C.R., H.I. conducted the mouse experiments. M.D. and A.P. supervised the analyses in the SUPER study. M.E.H. and S.S.G. supervised the analyses in the NFID study. C.-Y.C. and H.R. wrote the original manuscript. R.T., T.G., M.L., G.S.A., T.S., L.U., T.F., M.D., A.P., E.A.T., H.H., M.E.H., S.S.G., and T.L. critically revised the paper. All authors reviewed and approved the final version of the manuscript.

## Supplementary Materials

Supplementary Note Supplementary Figures S1 to S18 Supplementary Tables S1 to S20 References

