## Supplementary Materials for "The impact of rare protein coding genetic variation on adult cognitive function"

### Table of Contents

**Supplementary note** 3

- **Brief summary of cognitive function-associated genes identified** 3
- **Biogen Biobank team** 6
- **The SUPER-Finland study researchers** 6

**Supplementary Figures** 9

**Reference** 33

#### Supplementary note

##### *Brief summary of cognitive function-associated genes identified*

**ADGRB2.** *ADGRB2* (adhesion G protein-coupled receptor B2; also known as *BAI2*) encodes an adhesion G protein coupled receptor (GPCR) that is one of the main mediators of signal transduction in the central nervous system. ADGRB2 is considered as an orphan GPCR (oGPCR), for which endogenous ligands have not yet been identified [1]. *ADGRB2* is primarily expressed in the brain (neurons and astrocytes in hippocampus, amygdala and cerebral cortex) [2,3]. Variants near *ADGRB2* have been associated with educational attainment in a genome-wide association study [4], and also found associated with other traits such as body mass index [5], smoking, Intraocular pressure [6], or parental longevity [7].

**KDM5B.** *KDM5B* (lysine demethylase 5B; also known as *JARID1B* or *PLU1*) encodes a lysine-specific histone demethylase in the jumonji/ARID domain-containing family of histone demethylases. The encoded protein can demethylate tri-, di- and monomethylated lysine 4 of histone H3 (H3K4me1/2/3) [8–10], which is broadly associated with enhancers and promoters of actively transcribed genomic loci. Mutations in *KDM5B* are the cause for an autosomal-recessive intellectual disability syndrome [11] (OMIM # 618109) and have further been found associated with schizophrenia [12] and autism spectrum disorder [13,14] in sequencing studies, where disrupted neuronal differentiation was suggested as a potential mechanism. A search on GWAS Catalog (https://www.ebi.ac.uk/gwas/; accessed on Feb. 6, 2022) did not identify significant associations of *KDM5B* variants in previous GWAS. However, we note that the association of *KDM5B* with RT may be influenced by its association with reduced handgrip strength we observed in UKB, which might contribute to the epidemiological observation in UKB that hand grip strength and cognitive function share common mechanisms [15].

**GIGYF1.** *GIGYF1* (GRB10 interacting GYF protein 1) encodes an adaptor protein (a member of the gyf family) that binds growth factor receptor-bound 10 (GRB10), which in turn binds activated insulin receptors and insulin-like growth factor-1 (IGF-1) receptors [16,17]. By influencing the insulin and IGF-1 signaling pathway, *GIGYF1* plays a role in metabolic diseases and related anthropometric traits. For instance, significant associations were identified in previous GWAS for hemoglobin [18], total cholesterol, low density lipoprotein cholesterol, glucose and apolipoprotein B levels [19]. *GIGYF1* was also associated with mosaic loss of chromosome Y (LOY) [20] and metabolic diseases including glucose and HbA1c levels and type 2 diabetes [21] in previous exome sequencing studies.

**ANKRD12.** *ANKRD12* (ankyrin repeat domain 12; also known as *ANCO-2*) encodes a member of the ankyrin repeats-containing cofactor (ANCO) family. ANCOs are transcriptional co-regulators that interact with both co-activators and co-repressors [22]. *ANKRD12* interacts with the p160 co-activators (by recruiting HDACs [histone deacetylases]) and the co-activator ADA3 (alteration/deficiency in activation 3) [22,23]. *ANKRD12* was found to be associated with corpuscular measures in GWAS [5,24].

**SLC8A1.** *SLC8A1* (solute carrier family 8 member A1; also known as *NC*X1) encodes a bidirectional calcium transporter, the cardiac sarcolemmal Na(+)-Ca(2+) exchanger, which is the primary mechanism for cardiac myocyte returning to its resting state following excitation (through extrusion of calcium) and plays a critical role in cardiac contractility [25]. *SLC8A1* expression is enriched in human heart tissue. *SLC8A1* has been shown to be associated with bone mineral density [26], blood pressure[27], blood biomarkers (for example IGF-1 [19]), electrocardiographic traits (PR interval [28], QT interval [29], etc.) and hand grip strength [30] among others.

**RC3H2.** *RC3H2* (ring finger and CCCH-type domains 2) encodes roquin-2 that belongs to a family of highly conserved RNA-binding proteins (roquins) that regulate their target genes on the post-transcriptional level. Roquins contain a RING (Really Interesting New Gene)-type E3 ubiquitin ligase domain, followed by a ROQ domain and a CCCH-type ZnF domain [31–33]. Roquins play key roles in maintaining peripheral immunological tolerance and autoimmune diseases [34]. It has been shown that *RC3H2* (and *RC3H1*) restricts T-cell activation and costimulation via *ICOS* and *OX40* to prevent inappropriate Tfh cell differentiation [35]. Roquin-2 is widely expressed in all human tissues. *RC3H2* showed genome-wide significant association with insomnia [36] and HbA1c [5] in GWAS.

**CACNA1A.** *CACNA1A* (calcium voltage-gated channel subunit alpha1 A) encodes the alpha-1A subunit of the voltage-dependent calcium channels. It is primarily expressed in neuronal tissue. Mutations in *CACNA1A* are a cause for type 2 episodic ataxia (OMIM #108500), spinocerebellar ataxia 6 (OMIM #183086), developmental and epileptic encephalopathy 42 (OMIM #617106) and familial hemiplegic migraine (OMIM #141500). *CACNA1A* was implicated in a previous educational attainment GWAS[4], but the top associated SNP and LD peak do not fall into the *CACNA1A* gene region, but rather located in the intergenic region between *CACNA1A* and *RPL12P42*. Other GWAS associations for *CACNA1A* include depressive symptoms [37], age at first birth [38] and brain region volume [39].

**BCAS3.** *BCAS3* (BCAS3 microtubule associated cell migration factor) encodes a large, highly conserved cytoskeletal protein involved in human embryogenesis and tumor angiogenesis [40,41]. It has recently been shown that *BCAS3* loss-of-function variants can cause Hengel-Maroofian-Schols syndrome (HEMARS; OMIM # 619641), which is an autosomal recessive neurodevelopmental disorder characterized by severe global developmental delay starting from infancy or early childhood with facial dysmorphism and brain abnormalities [41]. *BCAS3* has also been associated with glomerular filtration rate [42], bone mineral density [26], serum creatinine level [5], hemoglobin concentration [24], serum urate level [43], red blood cell count [5], ophthalmologic measures (e.g. macular thickness [44]), coronary artery disease [45] and additional traits in GWAS.

***Biogen Biobank team***

Steering team
Ellen Tsai, Christopher D. Whelan, Paola Bronson, David Sexton, Sally John, Heiko Runz
Data management team
Eric Marshall, Mehool Patel, Saranya Duraisamy, Timothy Swan
Extended Scientific team
Dennis Baird, Chia-Yen Chen, Susan Eaton, Jake Gagnon, Feng Gao, Cynthia Gubbels, Yunfeng Huang, Varant Kupelian, Kejie Li, Dawei Liu, Stephanie Loomis, Helen McLaughlin, Adele Mitchell, Nilanjana Sadhu, Benjamin Sun, Ruoyu Tian

***The SUPER-Finland study researchers***

| | **AFFILIATION** |
|---|---|
| Aarno Palotie | Institute for Molecular Medicine, Finland (FIMM), HiLIFE, University of Helsinki, Helsinki, Finland; Broad Institute of MIT and Harvard, Cambridge, MA, USA; Massachusetts General Hospital Massachusetts General Hospital, Boston, MA, USA |
| Aija Kyttälä | Finnish Institute for Health and Welfare (THL), Helsinki, Finland |
| Amanda Elliott | Institute for Molecular Medicine Finland, HiLIFE, University of Helsinki, Finland; Broad Institute, Cambridge, MA, USA and Massachusetts General Hospital, Boston, MA, USA |
| Andre Sourander | Department of Child Psychiatry, University of Turku, Turku, Finland |
| Annamari Tuulio-Henriksson | Department of Psychology and Logopedics, Faculty of Medicine, University of Helsinki, Helsinki, Finland |
| Anssi Solismaa | Tampere University and Tampere University Hospital, Tampere, Finland |
| Antti Tanskanen | Department of Clinical Neuroscience, Karolinska Institutet, Stockholm, Sweden and Impact Assessment Unit, Finnish Institute for Health and Welfare (THL), Helsinki, Finland |
| Ari Ahola-Olli | Institute for Molecular Medicine Finland, HiLIFE, University of Helsinki, Helsinki, Finland |
| Arto Mustonen | University of Turku, Turku, Finland |
| Arttu Honkasalo | University of Helsinki, Helsinki, Finland |
| Asko Wegelius | Department of Psychiatry, University of Helsinki and Helsinki University Hospital, Finland |
| Atiqul Mazumder | Unit of Clinical Neuroscience, Faculty of Medicine, University of Oulu, Oulu, Finland |
| Auli Toivola | Finnish Institute for Health and Welfare (THL), Helsinki, Finland |
| Benjamin Neale | Broad Institute of MIT and Harvard, Cambridge, MA, USA |
| Elina Hietala | Tampere University and Tampere University Hospital |
| Elmo Saarentaus | Institute for Molecular Medicine Finland, HiLIFE, University of Helsinki, Helsinki, Finland |
| Erik Cederlöf | Finnish Institute for Health and Welfare (THL), Helsinki, Finland |
| Erkki Isometsä | Department of Psychiatry, University of Helsinki and Helsinki University Hospital, Helsinki, Finland |

| | |
|---|---|
| Heidi Taipale | Kuopio Research Center of Geriatric Care, University of Eastern Finland, Kuopio, Finland and School of Pharmacy, University of Eastern Finland, Kuopio, Finland, Department of Clinical Neuroscience, Karolinska Institutet, Stockholm, Sweden |
| Imre Västrik | Institute for Molecular Medicine Finland, HiLIFE, University of Helsinki, Helsinki, Finland |
| Jaana Suvisaari | Mental Health Unit, Finnish Institute for Health and Welfare, Helsinki, Finland |
| Jari Tiihonen | Department of Clinical Neuroscience, Karolinska Institutet, Stockholm, Sweden and Department of Forensic Psychiatry, Niuvanniemi Hospital, University of Eastern Finland, Kuopio, Finland |
| Jarmo Hietala | Department of Psychiatry, Turku University Hospital, Turku, Finland |
| Johan Ahti | Department of Psychiatry, University of Helsinki and Helsinki University Hospital, Helsinki, Finland |
| Jonne Lintunen | Department of Forensic Psychiatry, Niuvanniemi Hospital, University of Eastern Finland, Kuopio, Finland |
| Jouko Lönnqvist | Finnish Institute for Health and Welfare (THL), Helsinki, Finland and University of Helsinki, Helsinki, Finland |
| Juha Veijola | Department of Psychiatry, Research Unit of Clinical Neuroscience, University of Oulu, Oulu, Finland and Department of psychiatry, University Hospital of Oulu, Oulu, Finland |
| Julia Moghadampour | Tampere University and Tampere University Hospital, Helsinki, Finland |
| Jussi Niemi-Pynttäri | Department of Psychiatry, University of Helsinki and Helsinki University Hospital, Helsinki, Finland |
| Kaisla Lahdensuo | Mehiläinen, Helsinki, Finland |
| Katja Häkkinen | Department of Forensic Psychiatry, Niuvanniemi Hospital, University of Eastern Finland, Kuopio, Finland |
| Katriina Hakakari | Hospital District of Helsinki and Uusimaa, Helsinki, Finland |
| Kimmo Suokas | Tampere University Hospital, Tampere, Finland and Department of Psychiatry, Pirkanmaa Hospital District, Tampere, Finland |
| Lea Urpa | Institute for Molecular Medicine Finland, HiLIFE, University of Helsinki, Helsinki, Finland |
| Marjo Taivalantti | Research unit of clinical neuroscience, Faculty of medicine, University of Oulu, Oulu, Finland |
| Mark Daly | Institute for Molecular Medicine, Finland (FIMM), HiLIFE, University of Helsinki, Helsinki, Finland; Broad Institute of MIT and Harvard, Cambridge, MA, USA; Massachusetts General Hospital Massachusetts General Hospital, Boston, MA, USA |
| Markku Lähteenvuo | Department of Forensic Psychiatry, Niuvanniemi Hospital, University of Eastern Finland, Kuopio, Finland |
| Martta Kerkelä | Research Unit of Clinical Neuroscience, University of Oulu, Oulu, Finland |
| Minna Holm | Mental Health Unit, Finnish Institute for Health and Welfare, Helsinki, Finland |
| Nina Lindberg | Hospital District of Helsinki and Uusimaa, Helsinki, Finland |
| Noora Ristiluoma | Finnish Institute for Health and Welfare (THL), Helsinki, Finland |
| Olli Kampman | Tampere University and Tampere University Hospital, Tampere, Finland |
| Olli Pietiläinen | Neuroscience Center, HiLIFE, University of Helsinki, Helsinki, Finland |

| | |
|---|---|
| Risto Kajanne | Institute for Molecular Medicine Finland, HiLIFE, University of Helsinki, Helsinki, Finland |
| Sari Lång-Tonteri | Hospital District of Helsinki and Uusimaa, Helsinki, Finland |
| Solja Niemelä | Department of Psychiatry, University of Turku, Turku, Finland |
| Steven E. Hyman | Broad Institute of MIT and Harvard, Cambridge, MA, USA |
| Susanna Rask | Tampere University and Tampere University Hospital |
| Tarjinder Singh | Broad Institute of MIT and Harvard, Cambridge, MA, USA |
| Teemu Männynsalo | Department of Psychiatry, University of Helsinki and Helsinki University Hospital, Helsinki, Finland |
| Tiina Paunio | Mental Health Unit, Finnish Institute for Health and Welfare, Helsinki, Finland |
| Tuomas Jukuri | Department of Psychiatry, Oulu University Hospital, Oulu, Finland |
| Tuomo Kiiskinen | Institute for Molecular Medicine Finland, HiLIFE, University of Helsinki, Helsinki, Finland |
| Tuula Kieseppä | Hospital District of Helsinki and Uusimaa, Helsinki, Finland |
| Ville Mäkipelto | University of Helsinki, Helsinki, Finland |
| Willehard Haaki | Department of Psychiatry, University of Turku, Turku, Finland and Department of Psychiatry, Turku University Hospital, Turku, Finland |
| Zuzanna Misiewicz | Institute for Molecular Medicine Finland, HiLIFE, University of Helsinki, Helsinki, Finland |

#### Supplementary Figures

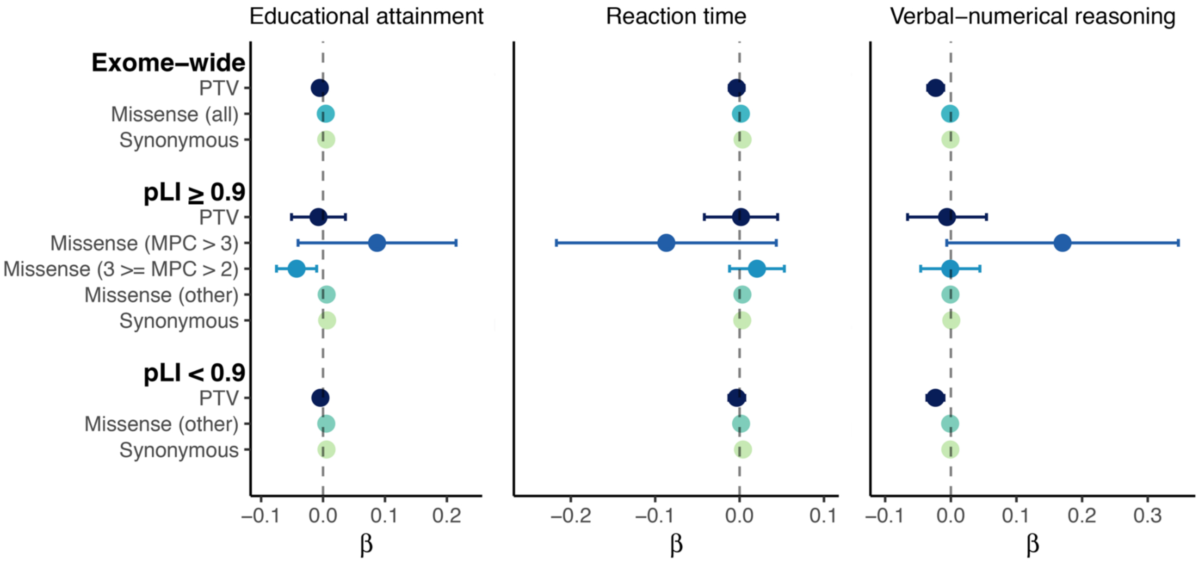


**Fig. S1. Impact of exome-wide burden of rare protein coding variants on educational attainment (EDU), reaction time (RT) and verbal-numerical reasoning (VNR) in South Asian (SAS) samples in the UK Biobank.**

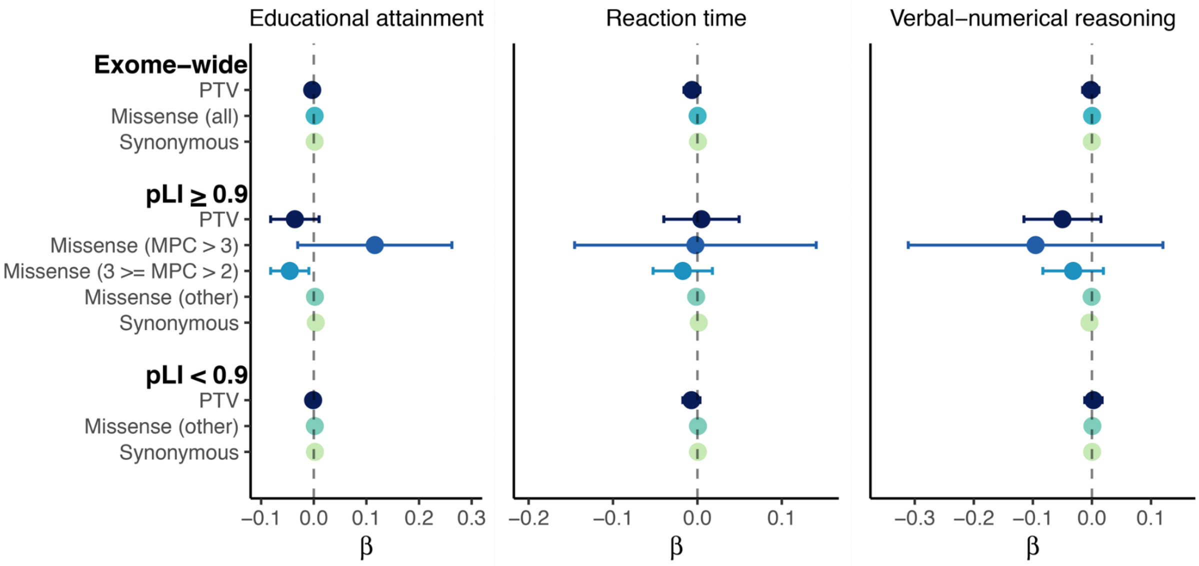


**Fig. S2. Impact of exome-wide burden of rare protein coding variants on educational attainment (EDU), reaction time (RT) and verbal-numerical reasoning (VNR) in African samples (AFR) in the UK Biobank.**

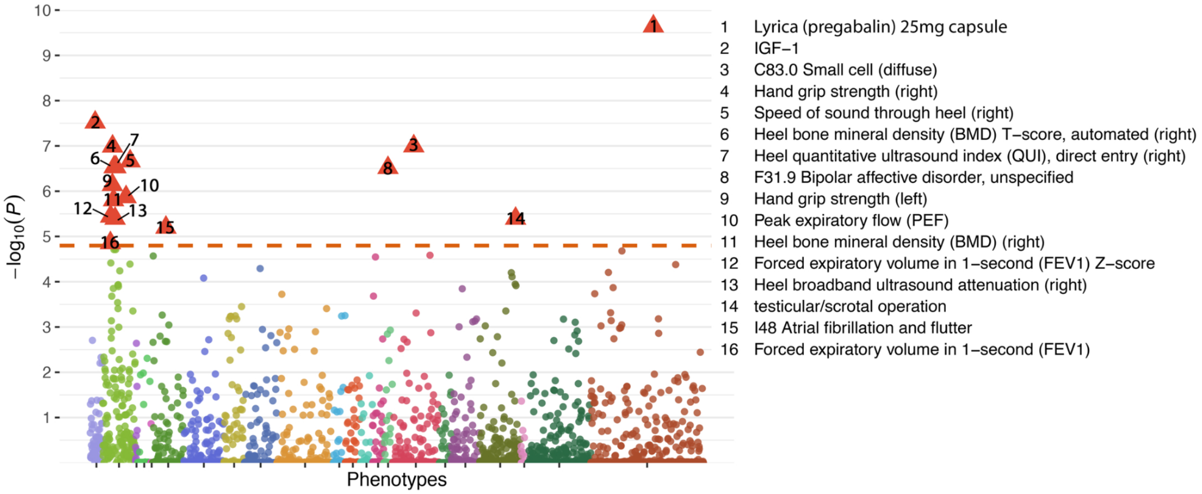


**Fig. S3. PTV burden-based phenome-wide association analysis (3,150 phenotypes) for KDM5B in the UK Biobank European samples.** FEV1 Z-score is Inverted GLI 2012 z-score for FEV1. Phenotypes were grouped and color-coded from left to right in the following categories: biomarker; composite phenotypes; family history; ICD-10 cause of death, ICD-10 congenital malformations; deformations and chromosomal abnormalities; ICD-10 diseases of the circulatory system; ICD-10 diseases of the digestive system; ICD-10 diseases of the eye and adnexa; ICD-10 diseases of the genitourinary system; ICD-10 diseases of the musculoskeletal system and connective tissue; ICD-10 diseases of the nervous system; ICD-10 diseases of the respiratory system; ICD-10 diseases of the skin and subcutaneous tissue; ICD-10 endocrine, nutritional and metabolic diseases; ICD-10 mental, behavioral and neurodevelopmental disorders; ICD-10 neoplasms; ICD-10 pregnancy, childbirth and the puerperium; ICD-10 symptoms, signs and abnormal clinical and laboratory findings, not elsewhere classified; operation code; self-reported illness: cancer; self-reported illness: non−cancer; self-reported medication.

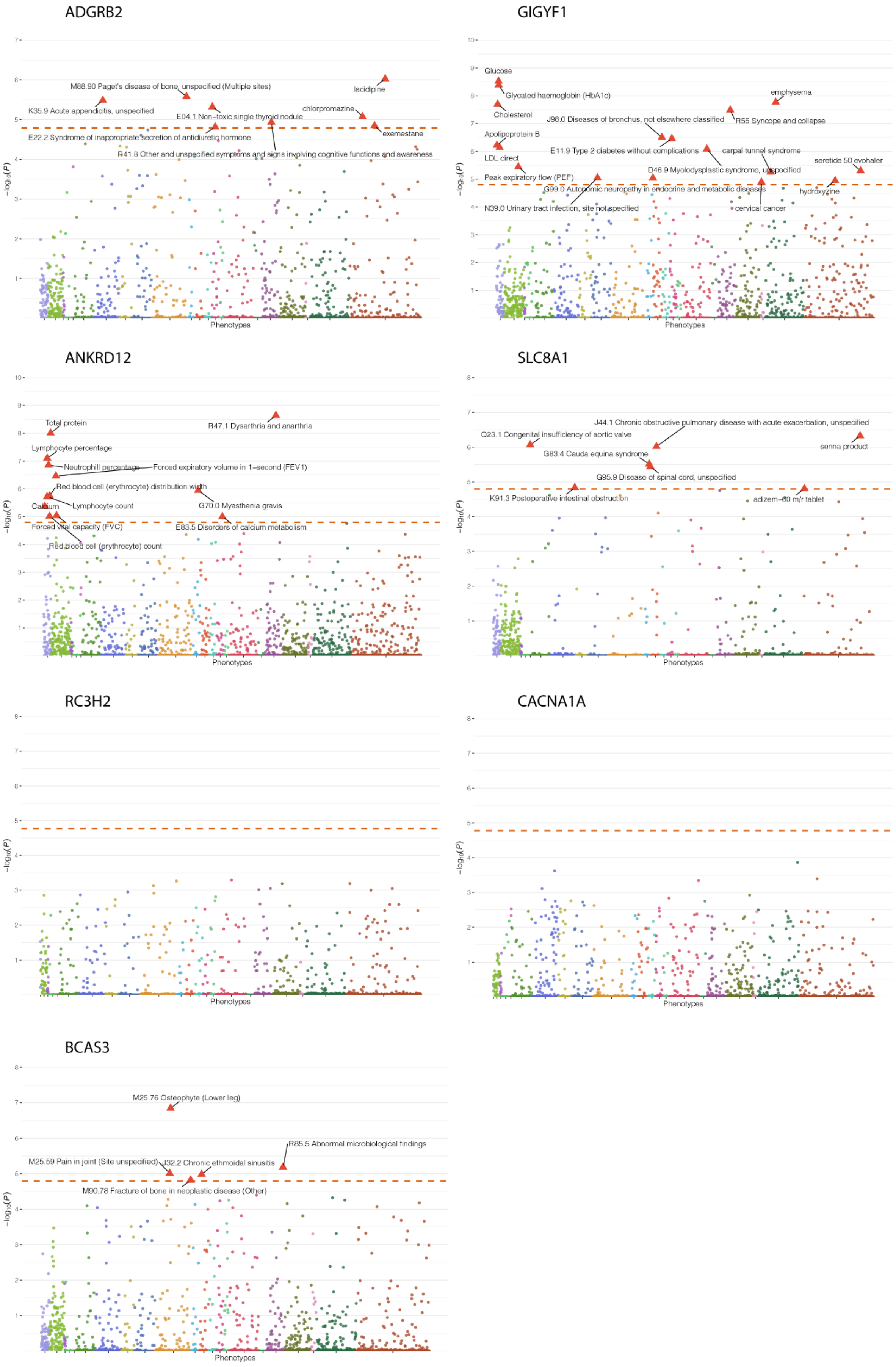

ADGRB2
M88.90 Paget's disease of bone, unspecified (Multiple sites)
lacidipine
K35.9 Acute appendicitis, unspecified
E04.1 Non-toxic single thyroid nodule
chlorpromazine
E22.2 Syndrome of inappropriate secretion of antidiuretic hormone
exemestane
R41.8 Other and unspecified symptoms and signs involving cognitive functions and awareness
Phenotypes
GIGYF1
Glucose
Glycated haemoglobin (HbA1c)
emphysema
Cholesterol
J98.0 Diseases of bronchus, not elsewhere classified
R55 Syncope and collapse
Apolipoprotein B
E11.9 Type 2 diabetes without complications
carpal tunnel syndrome
LDL direct
seretide 50 evohaler
D46.9 Myelodysplastic syndrome, unspecified
Peak expiratory flow (PEF)
G99.0 Autonomic neuropathy in endocrine and metabolic diseases
hydroxyzine
N39.0 Urinary tract infection, site not specified
cervical cancer
Phenotypes
ANKRD12
Total protein
R47.1 Dysarthria and anarthria
Lymphocyte percentage
Neutrophill percentage
Forced expiratory volume in 1-second (FEV1)
Red blood cell (erythrocyte) distribution width
Calcium
Lymphocyte count
G70.0 Myasthenia gravis
Forced vital capacity (FVC)
E83.5 Disorders of calcium metabolism
Red blood cell (erythrocyte) count
Phenotypes
SLC8A1
J44.1 Chronic obstructive pulmonary disease with acute exacerbation, unspecified
Q23.1 Congenital insufficiency of aortic valve
senna product
G83.4 Cauda equina syndrome
G95.9 Disease of spinal cord, unspecified
K91.3 Postoperative intestinal obstruction
adizem-60 m/r tablet
Phenotypes
RC3H2
Phenotypes
CACNA1A
Phenotypes
BCAS3
M25.76 Osteophyte (Lower leg)
R85.5 Abnormal microbiological findings
M25.59 Pain in joint (Site unspecified)
J32.2 Chronic ethmoidal sinusitis
M90.78 Fracture of bone in neoplastic disease (Other)
Phenotypes
$-\log_{10}(P)$

**Fig. S4. Phenome-wide association analysis (3,150 phenotypes) for *ADGRB2*, *GIGYF1*, *ANKRD12*, *SLC8A1*, *RC3H2, CACNA1A* and *BCAS3* in unrelated European samples in the UK Biobank.** Phenotypes were grouped and color-coded from left to right in the following categories: biobmarker; composite phenotypes; family history; ICD-10 cause of death, ICD-10 congenital malformations; deformations and chromosomal abnormalities; ICD-10 diseases of the circulatory system; ICD-10 diseases of the digestive system; ICD-10 diseases of the eye and adnexa; ICD-10 diseases of the genitourinary system; ICD-10 diseases of the musculoskeletal system and connective tissue; ICD-10 diseases of the nervous system; ICD-10 diseases of the respiratory system; ICD-10 diseases of the skin and subcutaneous tissue; ICD-10 endocrine, nutritional and metabolic diseases; ICD-10 mental, behavioral and neurodevelopmental disorders; ICD-10 neoplasms; ICD-10 pregnancy, childbirth and the puerperium; ICD-10 symptoms, signs and abnormal clinical and laboratory findings, not elsewhere classified; operation code; self-reported illness: cancer; self-reported illness: non−cancer; self-reported medication.

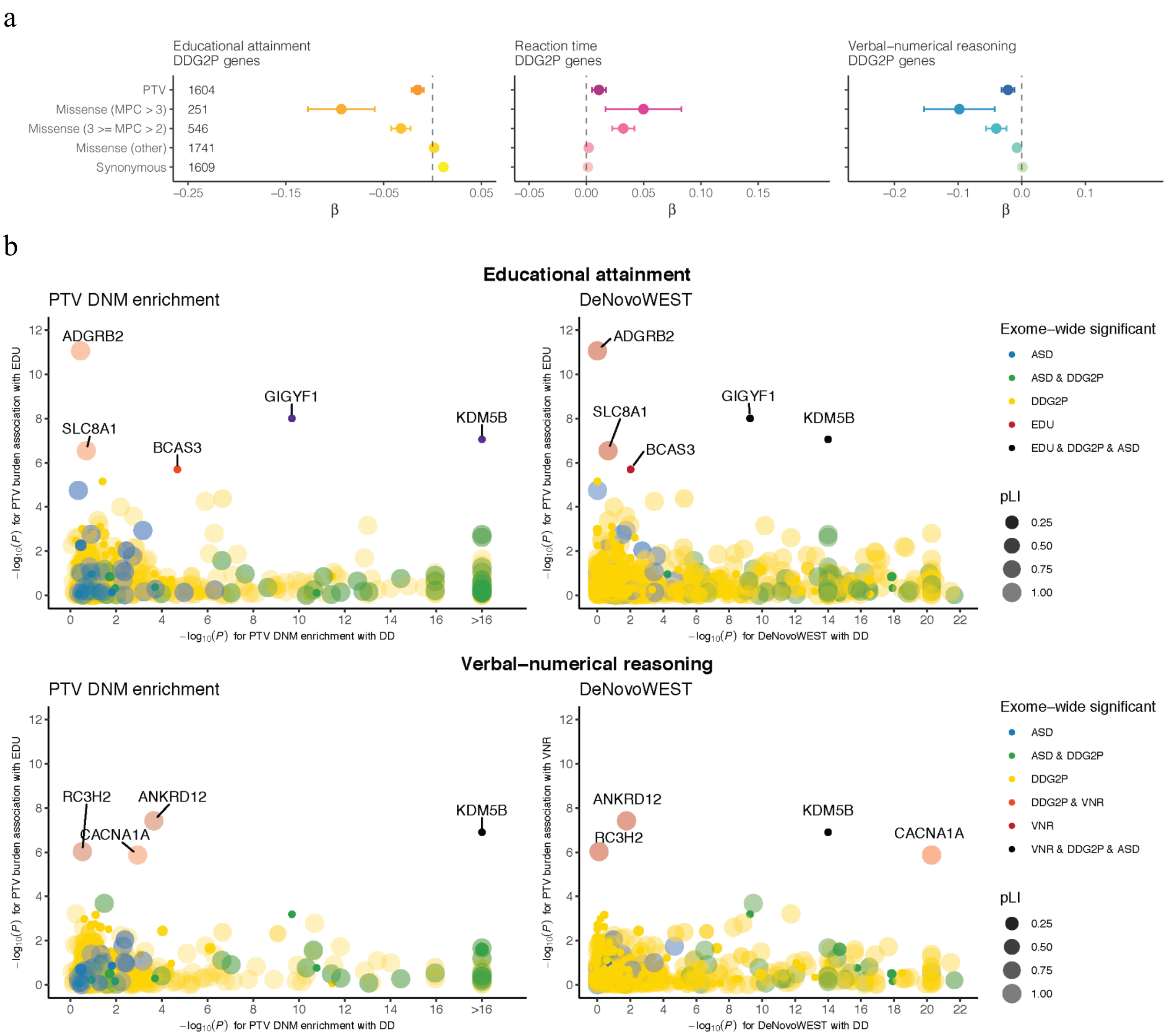


**Fig. S5. Impact of rare coding variants in genes identified in the Developmental Disorder Genotype - Phenotype Database (DDG2P) on cognitive function.**

**a.** The effects of protein-truncating, missense (stratified by MPC) and synonymous variant burden in exome sequencing study identified DDG2P on EDU, RT and VNR. DDG2P database (https://www.deciphergenomics.org/ddd/ddgenes) was accessed on December 23, 2020. Missense variants were classified by deleteriousness (MPC) into 3 tiers: tier 1 with MPC > 3; tier 2 with 3 ≥ MPC > 2; tier 3 includes all missense variants not in tier 1 or 2. We note that the effect of damaging missense variants out scaled that of PTV burden for DDG2P genes. This is most likely explained by UKB participants being depleted for highly penetrant PTVs in this gene set that cause disease onset in childhood[46].

**b.** Comparison between gene-based associations for genes from DDG2P database, EDU and VNR (PTV DNM enrichment and DeNovoWEST for DD; rare PTV burden for EDU and VNR). Each dot represents a gene that is identified for DD in Kaplanis et al. 2020 and for EDU or VNR in the current exome analysis. The dots are color-coded according to the phenotypes (DD, ASD, or EDU) that the gene is exome-wide significantly associated with. The size and shade of the dots are representing the pLI for the gene. EDU and VNR genes are labeled with gene names.

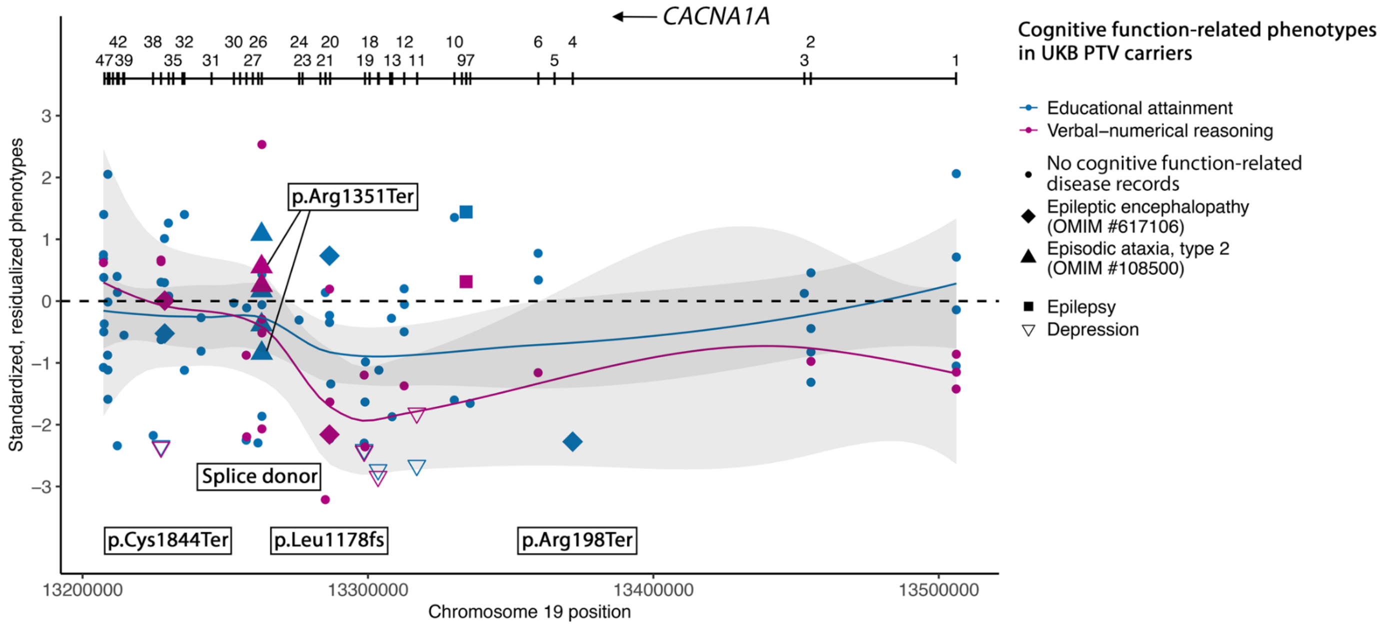


**Fig. S6. Distribution of cognitive phenotypes (educational attainment and verbal-numerical reasoning) for *CACNA1A* PTV carriers.** ClinVar pathogenic/likely pathogenic variants for epileptic encephalopathy (OMIM #617106) and/or type 2 episodic ataxia (OMIM #108500) was annotated. Samples with inpatient ICD-10 (International Classification of Diseases version-10) records of psychiatric (schizophrenia, bipolar disorder, depression, substance use disorder and/or anxiety and stress disorders), neurodegenerative and neurodevelopmental disorders were annotated. Phenotypes were residualized by sex, age, age$^2$, sex by age interaction, sex by age$^2$ interaction, top 20 PCs, and recruitment center and inverse rank-based normal transformed.

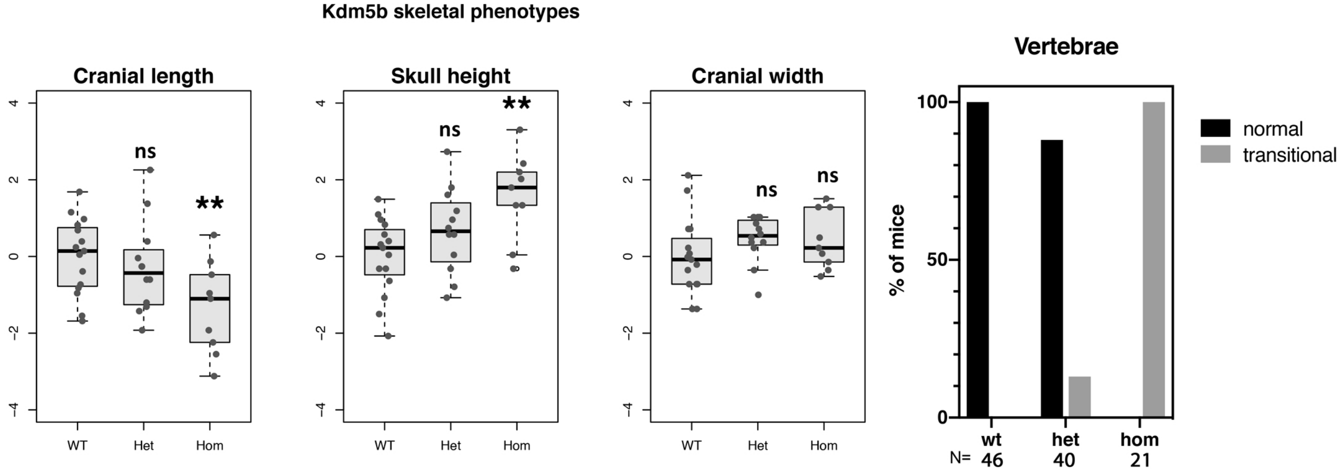


**Fig. S7. *Kdm5b* loss-of-function impacts craniofacial and skeletal features in mice in a dose-dependent manner.** An intermediate effect on cranial length (additive genotype effect *P*=0.0268) and height (additive genotype effect *P*=0.0056) is detected in $Kdm5b^{+/-}$ mice, but not in cranial width (additive genotype effect *P*=0.3090). A fully penetrant transitional vertebrae phenotype seen in $Kdm5b^{-/-}$ mice (N=21, Fisher's exact test vs $Kdm5b^{+/+}$ [N=46] *P*<0.001) is observed at a lower frequency in $Kdm5b^{+/-}$ mice (N=40, Fisher's exact test vs $Kdm5b^{+/+}$ *P*=0.0189).

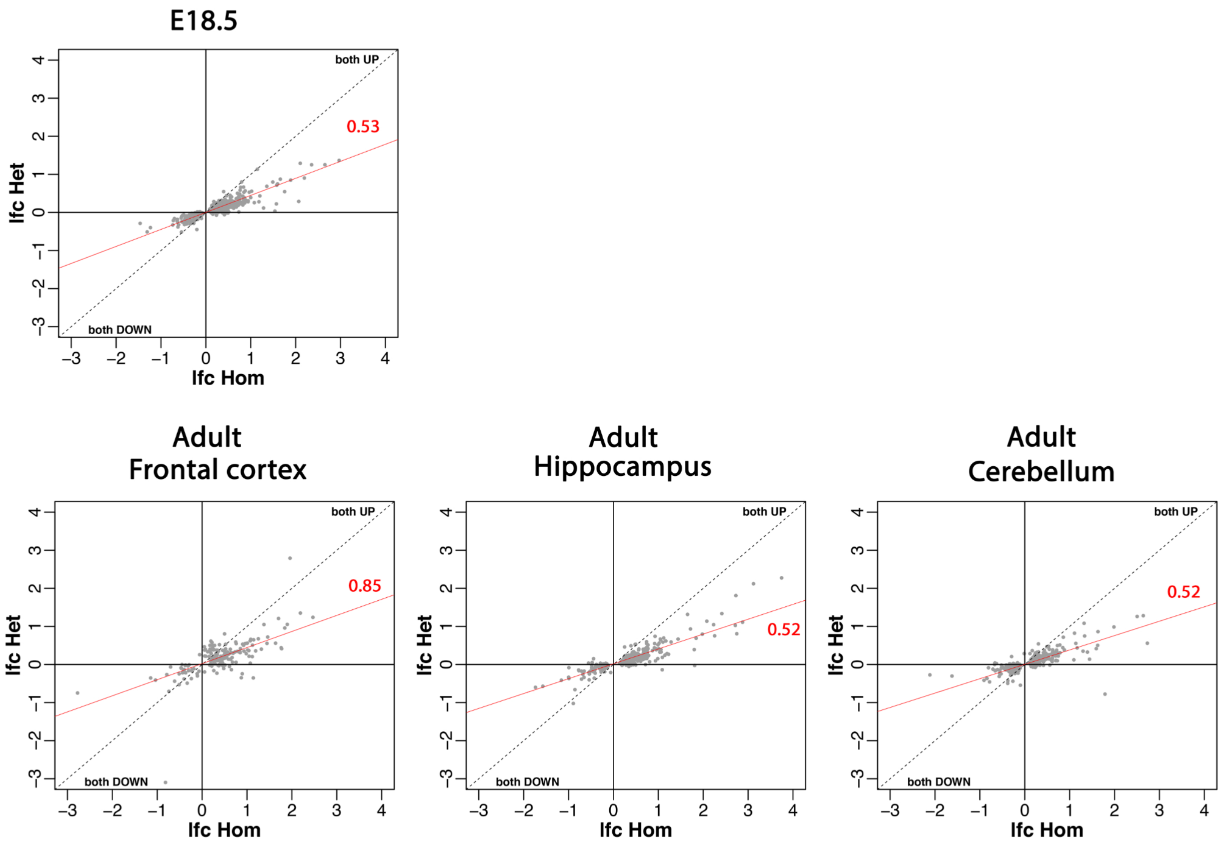


**Fig. S8. Correlation in differential gene expression between heterozygous and homozygous *Kdm5b* mutant mice.** LOG2-fold change of differentially expressed genes plotted for *Kdm5b*$^{+/-}$ (y-axis) and *Kdm5b*$^{-/-}$ (x-axis) mice across embryonic and adult brain tissues as indicated. There is a strong correlation between direction of change in expression in both mutant genotypes (robust linear regression line and slope shown in red).

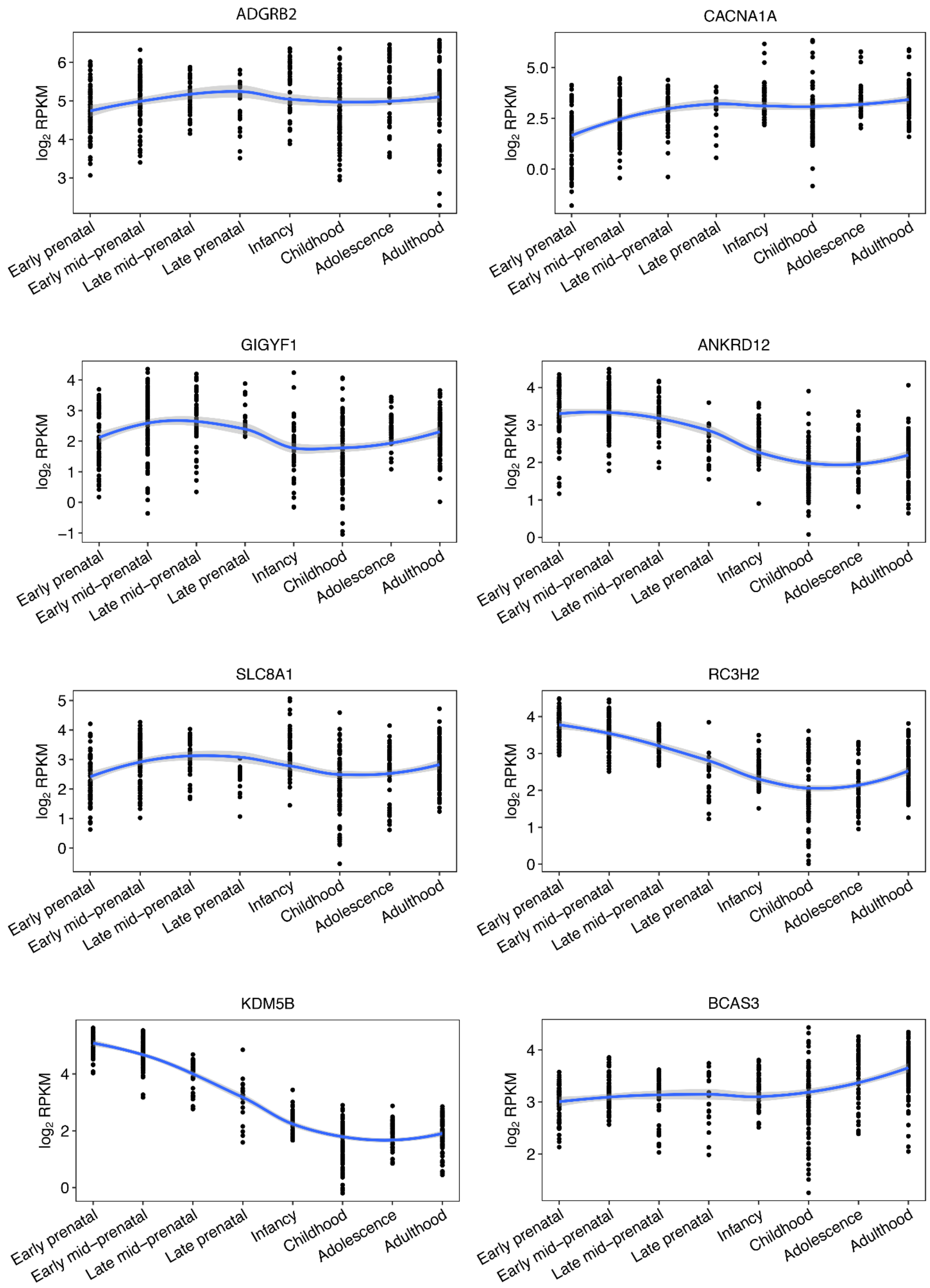


**Fig. S9. Cognitive function gene expression in brain tissue at different developmental stages. RNA-seq data obtained from BrainSpan[47].** Blue line represents fitted loess regression on *KDM5B* expression cross development stages.

**E18.5**

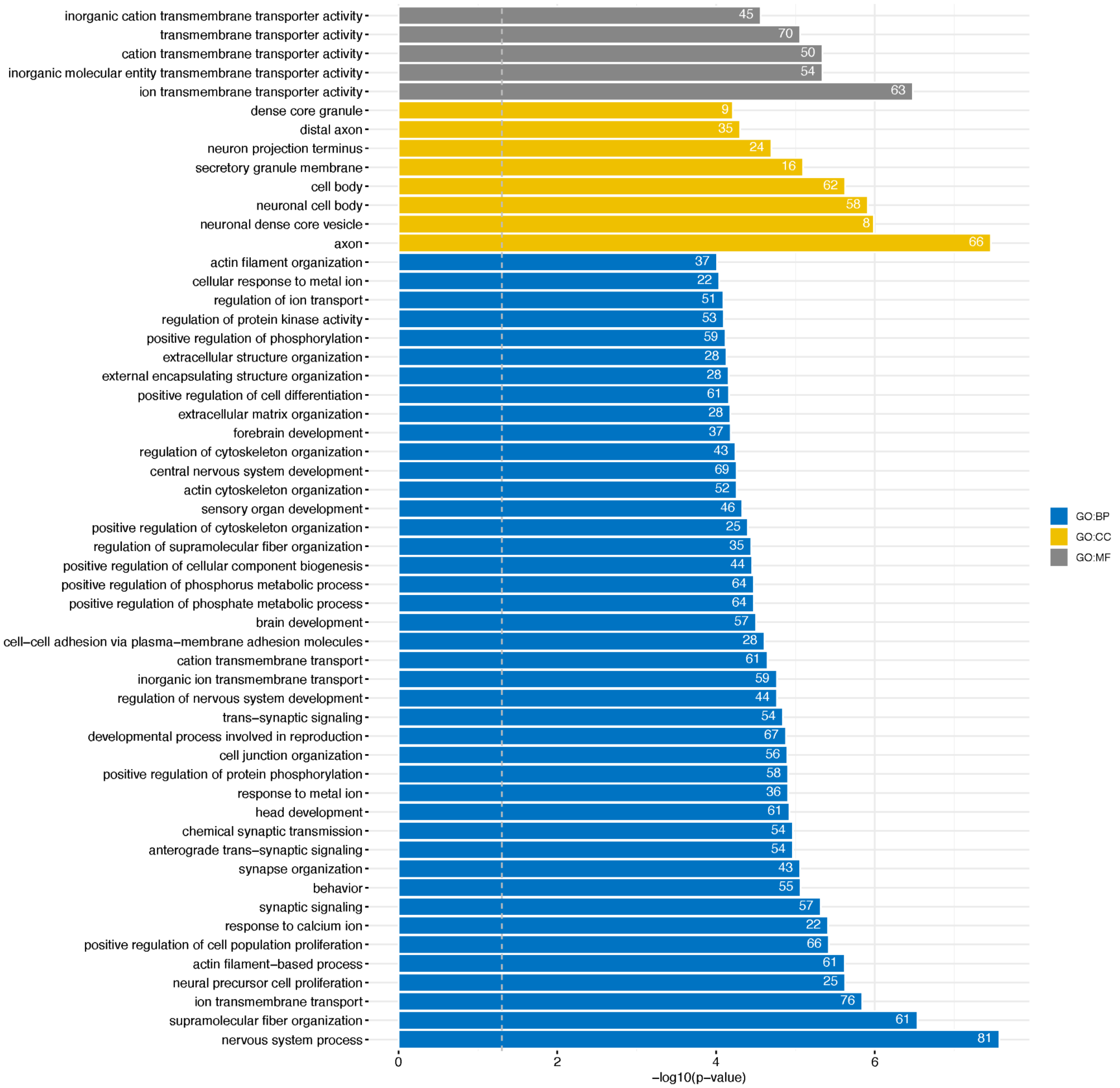

inorganic cation transmembrane transporter activity
transmembrane transporter activity
cation transmembrane transporter activity
inorganic molecular entity transmembrane transporter activity
ion transmembrane transporter activity
dense core granule
distal axon
neuron projection terminus
secretory granule membrane
cell body
neuronal cell body
neuronal dense core vesicle
axon
actin filament organization
cellular response to metal ion
regulation of ion transport
regulation of protein kinase activity
positive regulation of phosphorylation
extracellular structure organization
external encapsulating structure organization
positive regulation of cell differentiation
extracellular matrix organization
forebrain development
regulation of cytoskeleton organization
central nervous system development
actin cytoskeleton organization
sensory organ development
positive regulation of cytoskeleton organization
regulation of supramolecular fiber organization
positive regulation of cellular component biogenesis
positive regulation of phosphorus metabolic process
positive regulation of phosphate metabolic process
brain development
cell−cell adhesion via plasma−membrane adhesion molecules
cation transmembrane transport
inorganic ion transmembrane transport
regulation of nervous system development
trans−synaptic signaling
developmental process involved in reproduction
cell junction organization
positive regulation of protein phosphorylation
response to metal ion
head development
chemical synaptic transmission
anterograde trans−synaptic signaling
synapse organization
behavior
synaptic signaling
response to calcium ion
positive regulation of cell population proliferation
actin filament−based process
neural precursor cell proliferation
ion transmembrane transport
supramolecular fiber organization
nervous system process
45
70
50
54
63
9
35
24
16
62
58
8
66
37
22
51
53
59
28
28
61
28
37
43
69
52
46
25
35
44
64
64
57
28
61
59
44
54
67
56
58
36
61
54
54
43
55
57
22
66
61
25
76
61
81
0
2
4
6
−log10(p−value)
GO:BP
GO:CC
GO:MF

### Adult Frontal Cortex

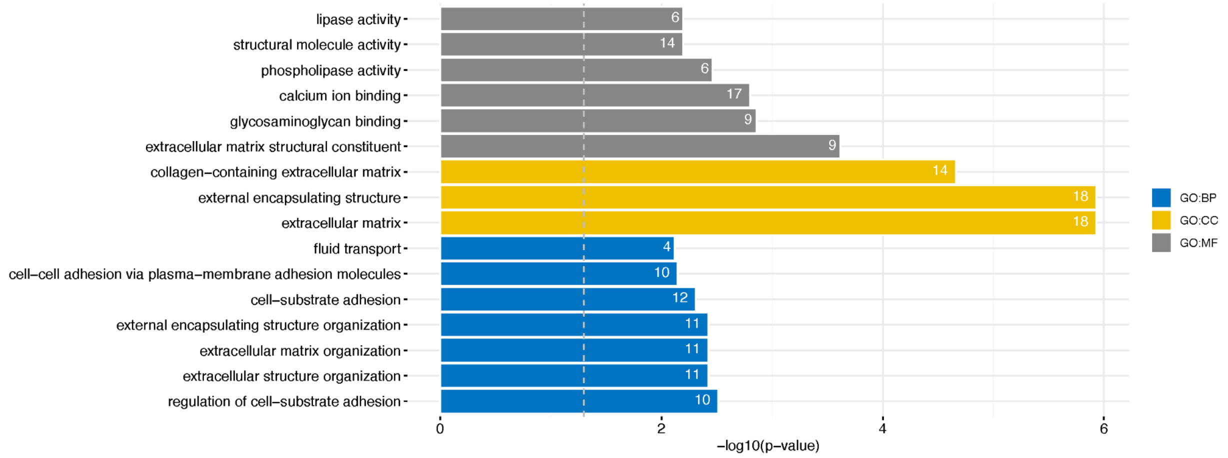


### Adult Hippocampus

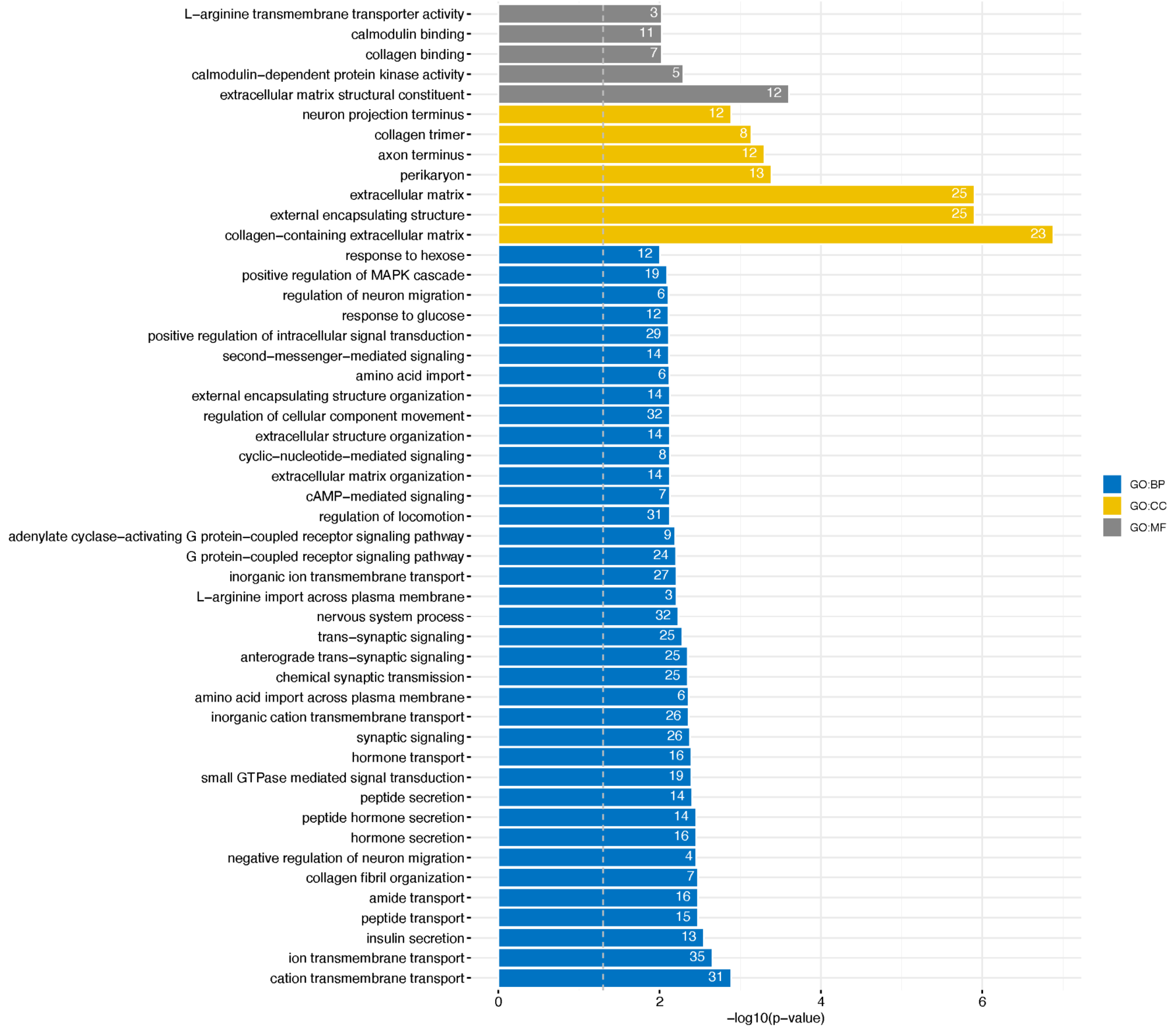

**Adult Cerebellum**

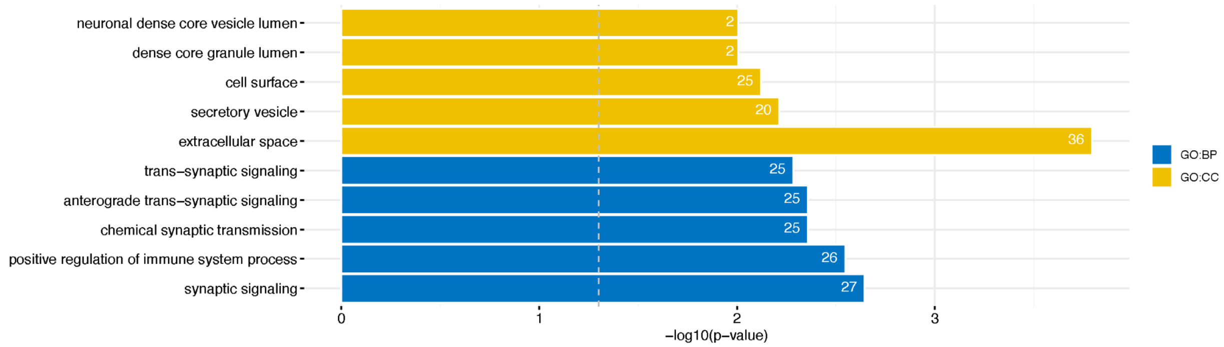


**Fig. S10. GO term enrichment for differentially expressed genes in *Kdm5b* mutant mice.** Differentially expressed genes (DEGs) from E18.5 and adult brain tissues of *Kdm5b*$^{+/-}$ and *Kdm5b*$^{-/-}$ mice were subject to Gene ontology (GO) pathway enrichment analysis using the gprofiler R package, with a threshold of 5% FDR and an enrichment significance threshold of $P$<0.05 (hypergeometric test with FDR correction for multiple testing). For the E18.5 sample, we only showed results with enrichment p-value<0.0001 (for display purposes). Full results are provided in Table S14. The European Nucleotide Archive accession numbers for the RNA-seq sequences reported are provided in Table S15. Background comprised only expressed genes in each tissue of interest.

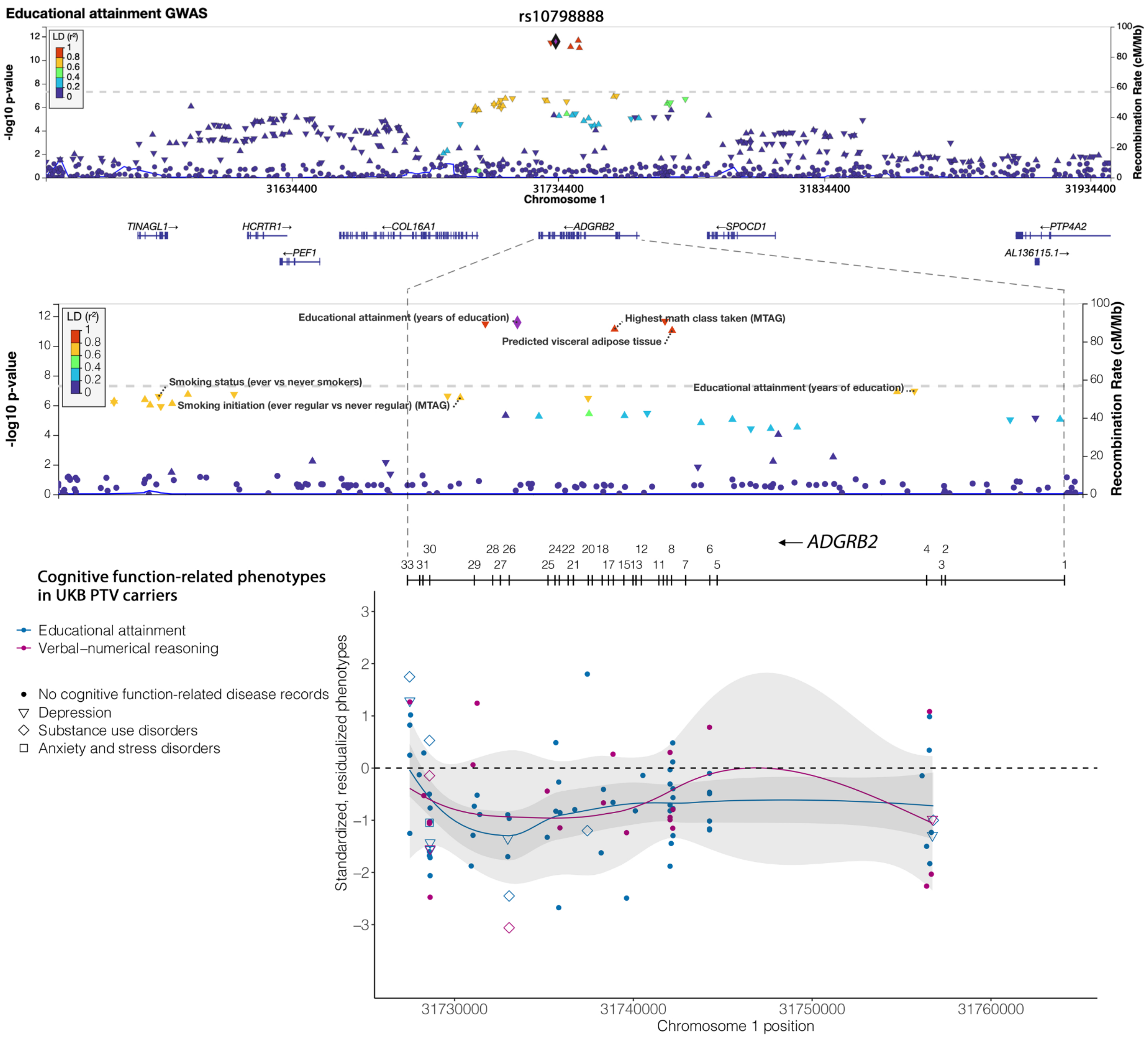


**Fig. S11. Overlap between educational attainment GWAS (Lee et al. 2018[4]) locus on chromosome 1 and *ADGBR2* identified in PTV burden analysis in UKB.** Regional plot of educational attainment GWAS association test results were generated around top independent SNP rs10798888. Additional associations from GWAS catalog were annotated with the associated phenotypes in the regional plot. EDU and VNR score for *ADGRB2* PTV carriers in UKB were plotted (both phenotypes were residualized by sex, age, age$^2$, sex by age, sex by age$^2$, top 20 PCs and recruitment centers and were inverse rank-based normal transformed). Samples with inpatient ICD-10 (International Classification of Diseases version-10) records of psychiatric, neurodegenerative, and neurodevelopmental disorders were annotated.

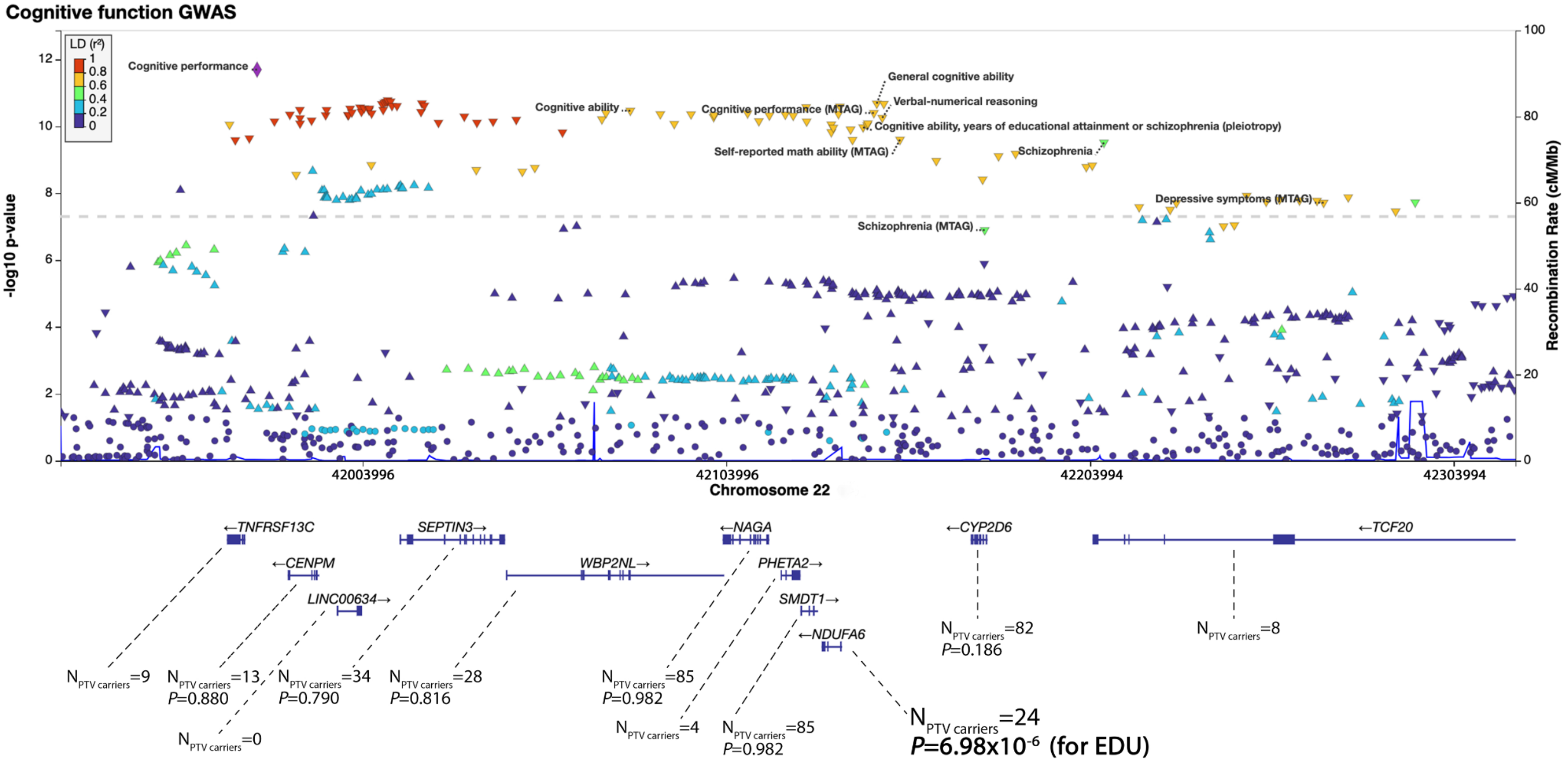


**Fig. S12. Overlap between cognitive function GWAS (Lam et al. 2021 [48]) locus on chromosome 22 and *NDUFA6* identified in PTV burden analysis in UKB (FDR significant for EDU).** Regional plot of cognitive function GWAS association test results were generated for top independent SNP rs5751191 and the extended LD region. Additional associations from GWAS catalog were annotated with the associated phenotypes in the regional plot. Number of PTV carriers and gene-based PTV burden association p-value were extracted for genes in the region.

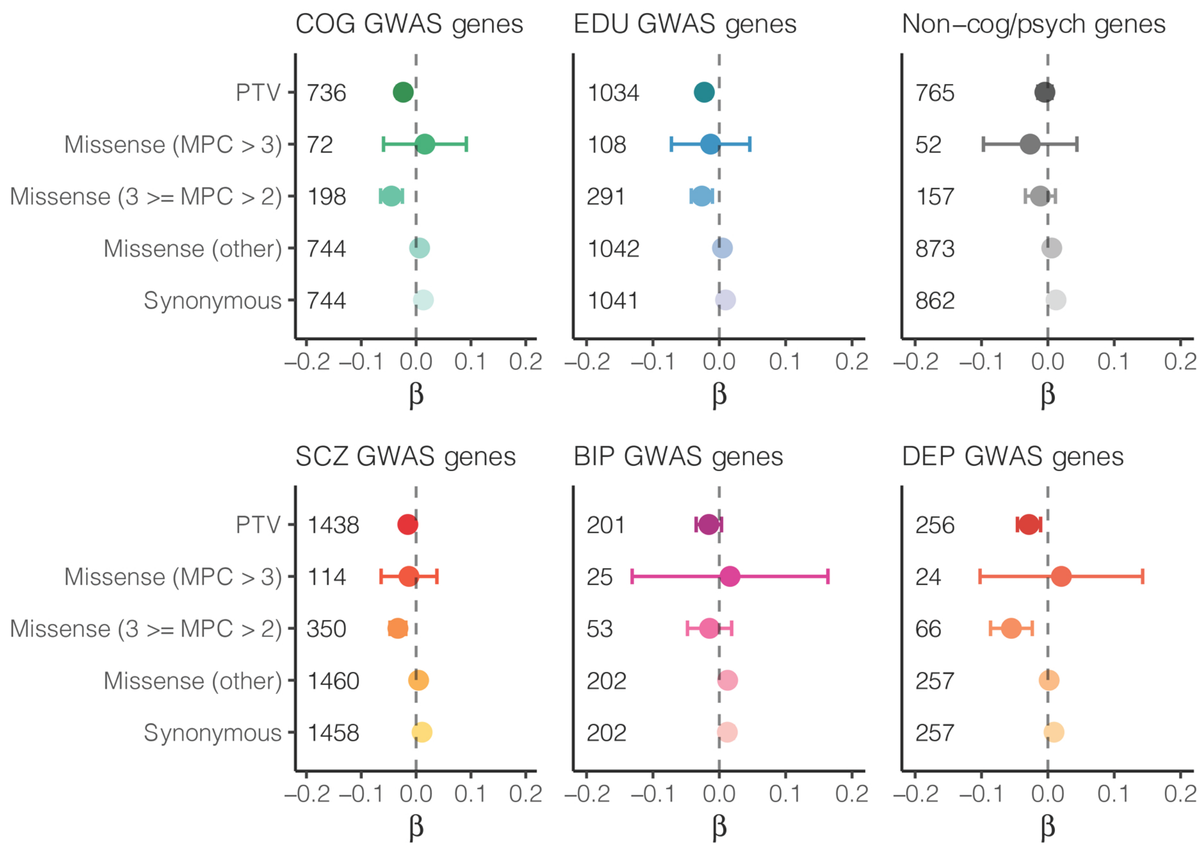


**Fig. S13. Rare coding variant burden in genes identified in GWAS for cognitive function, educational attainment, schizophrenia, bipolar disorder and depression and non-cognitive function related genes on educational attainment (EDU).** The impact of rare coding variant burden in genes identified through common variant association in GWAS for cognitive function (COG), educational attainment (EDU), schizophrenia (SCZ), bipolar disorder (BIP) and depression (DEP) and in non-cognitive function/non-psychiatric disorder-related (non-cog/psych) genes on EDU. Missense variants were classified by deleteriousness (MPC) into 3 tiers: MPC>3; 3≥MPC>2; and all missense variants not in the previous two tiers. The number of genes included in each burden was labeled in each panel.

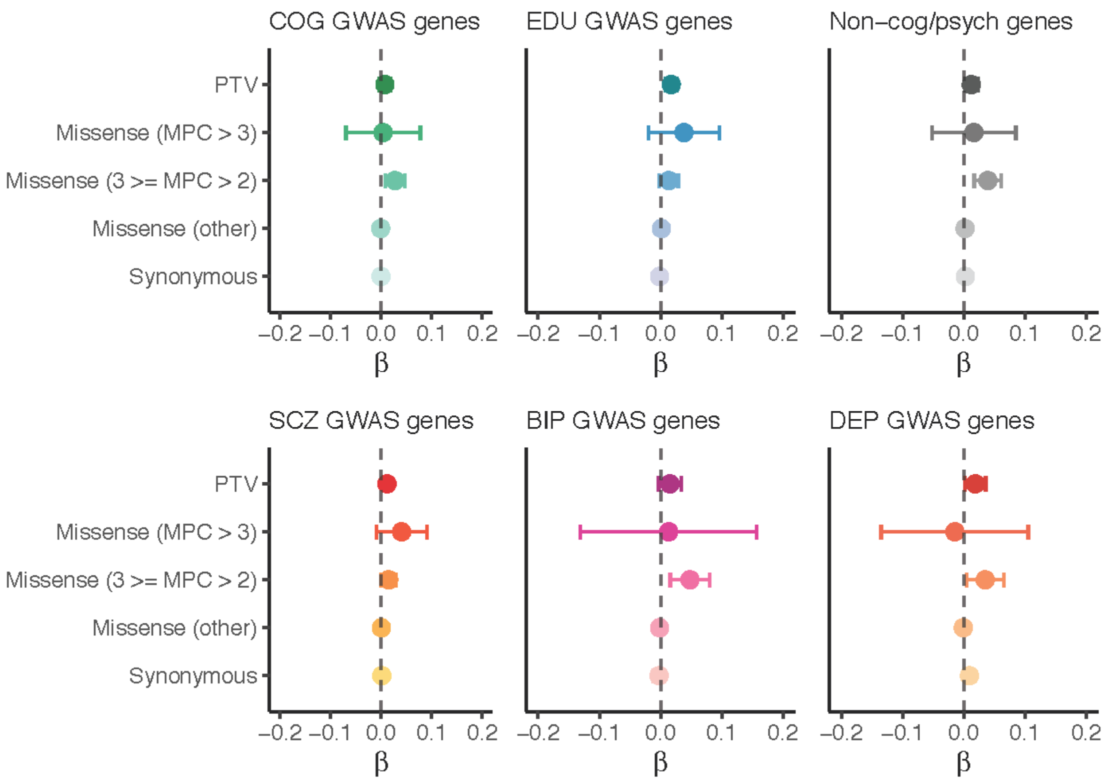


**Fig. S14. Rare coding variant burden in genes identified in GWAS for cognitive function, educational attainment, schizophrenia, bipolar disorder and depression and non-cognitive function related genes on reaction time (RT).**

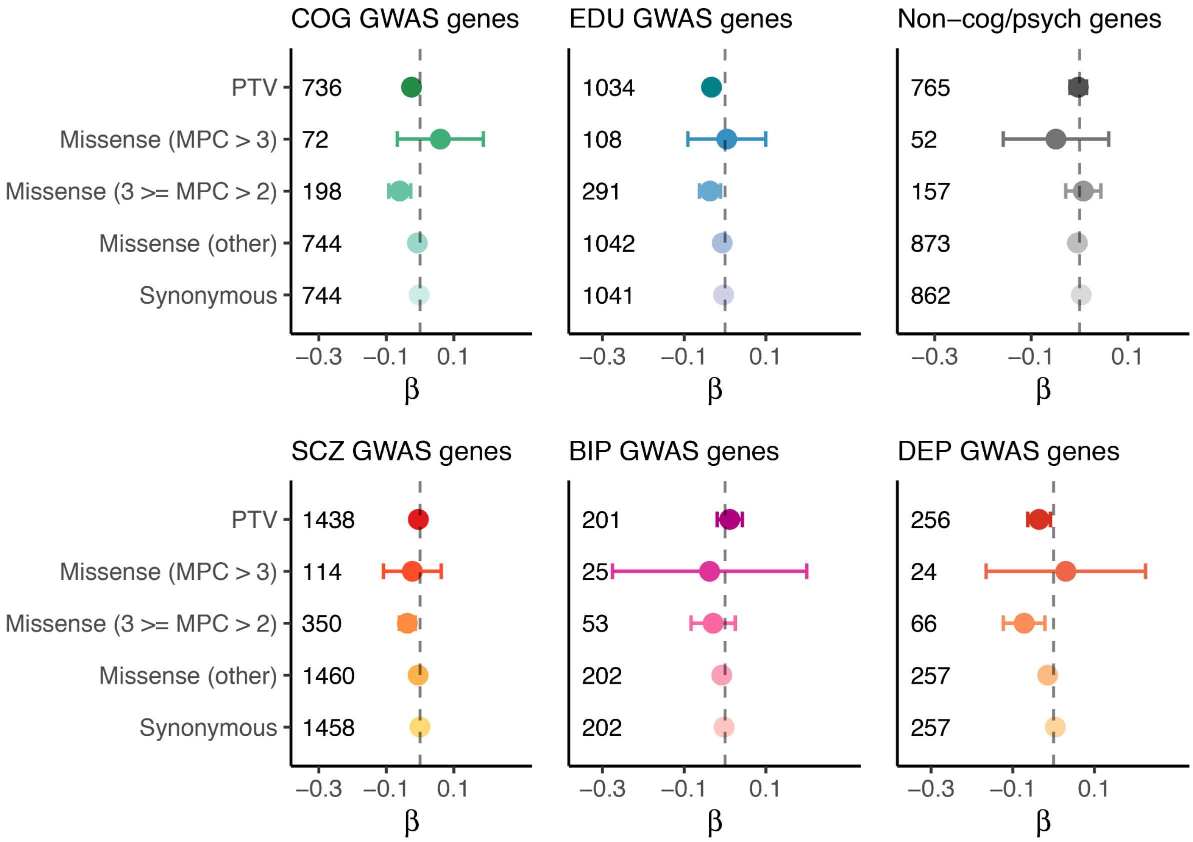


**Fig. 15. The impact of rare coding variant burdens in genes identified in GWAS for cognitive function, educational attainment, schizophrenia, bipolar disorder and depression and non-cognitive function related genes on verbal-numerical reasoning (VNR).**

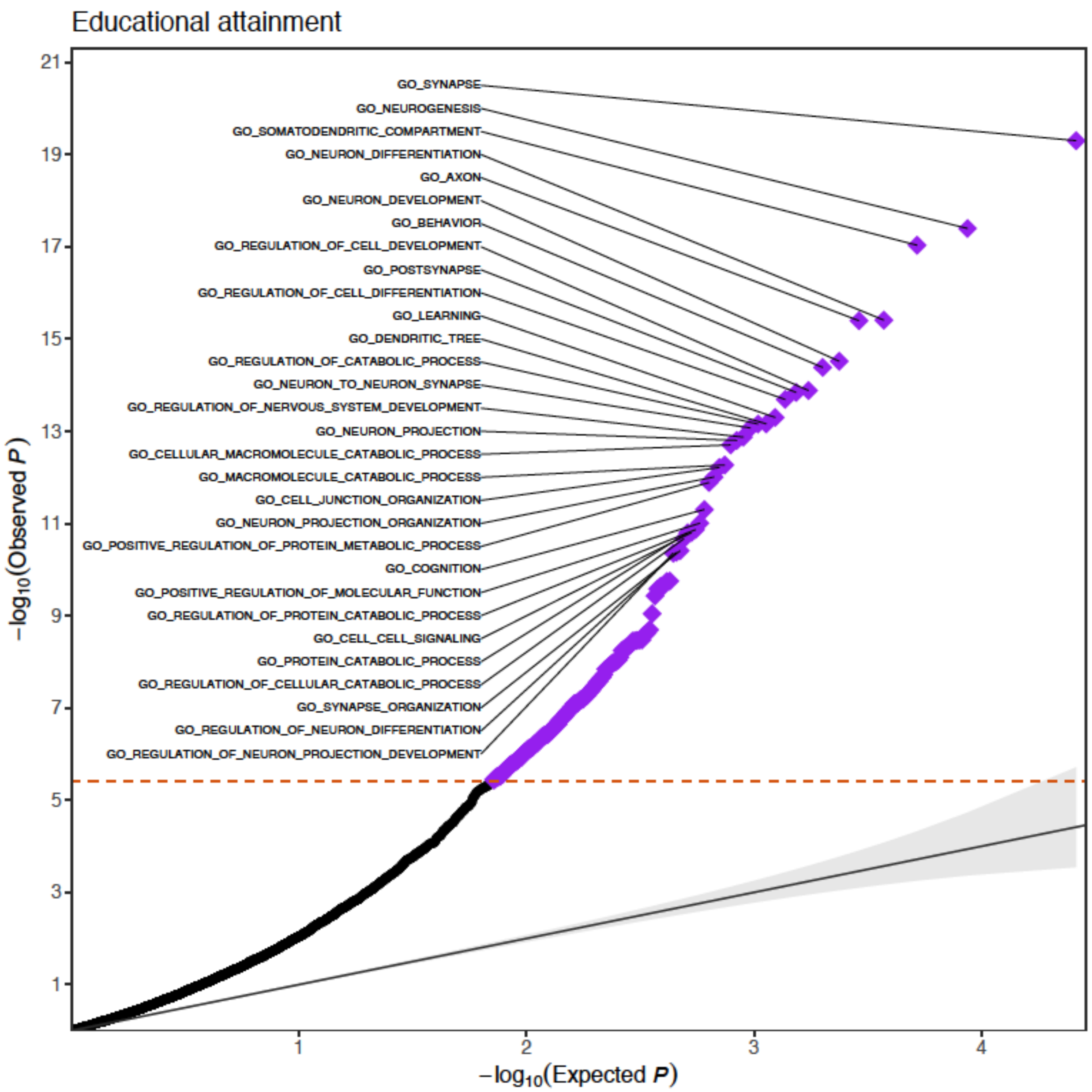

Educational attainment
GO_SYNAPSE
GO_NEUROGENESIS
GO_SOMATODENDRITIC_COMPARTMENT
GO_NEURON_DIFFERENTIATION
GO_AXON
GO_NEURON_DEVELOPMENT
GO_BEHAVIOR
GO_REGULATION_OF_CELL_DEVELOPMENT
GO_POSTSYNAPSE
GO_REGULATION_OF_CELL_DIFFERENTIATION
GO_LEARNING
GO_DENDRITIC_TREE
GO_REGULATION_OF_CATABOLIC_PROCESS
GO_NEURON_TO_NEURON_SYNAPSE
GO_REGULATION_OF_NERVOUS_SYSTEM_DEVELOPMENT
GO_NEURON_PROJECTION
GO_CELLULAR_MACROMOLECULE_CATABOLIC_PROCESS
GO_MACROMOLECULE_CATABOLIC_PROCESS
GO_CELL_JUNCTION_ORGANIZATION
GO_NEURON_PROJECTION_ORGANIZATION
GO_POSITIVE_REGULATION_OF_PROTEIN_METABOLIC_PROCESS
GO_COGNITION
GO_POSITIVE_REGULATION_OF_MOLECULAR_FUNCTION
GO_REGULATION_OF_PROTEIN_CATABOLIC_PROCESS
GO_CELL_CELL_SIGNALING
GO_PROTEIN_CATABOLIC_PROCESS
GO_REGULATION_OF_CELLULAR_CATABOLIC_PROCESS
GO_SYNAPSE_ORGANIZATION
GO_REGULATION_OF_NEURON_DIFFERENTIATION
GO_REGULATION_OF_NEURON_PROJECTION_DEVELOPMENT
$-\log_{10}$(Observed $P$)
21
19
17
15
13
11
9
7
5
3
1
1
2
3
4
$-\log_{10}$(Expected $P$)

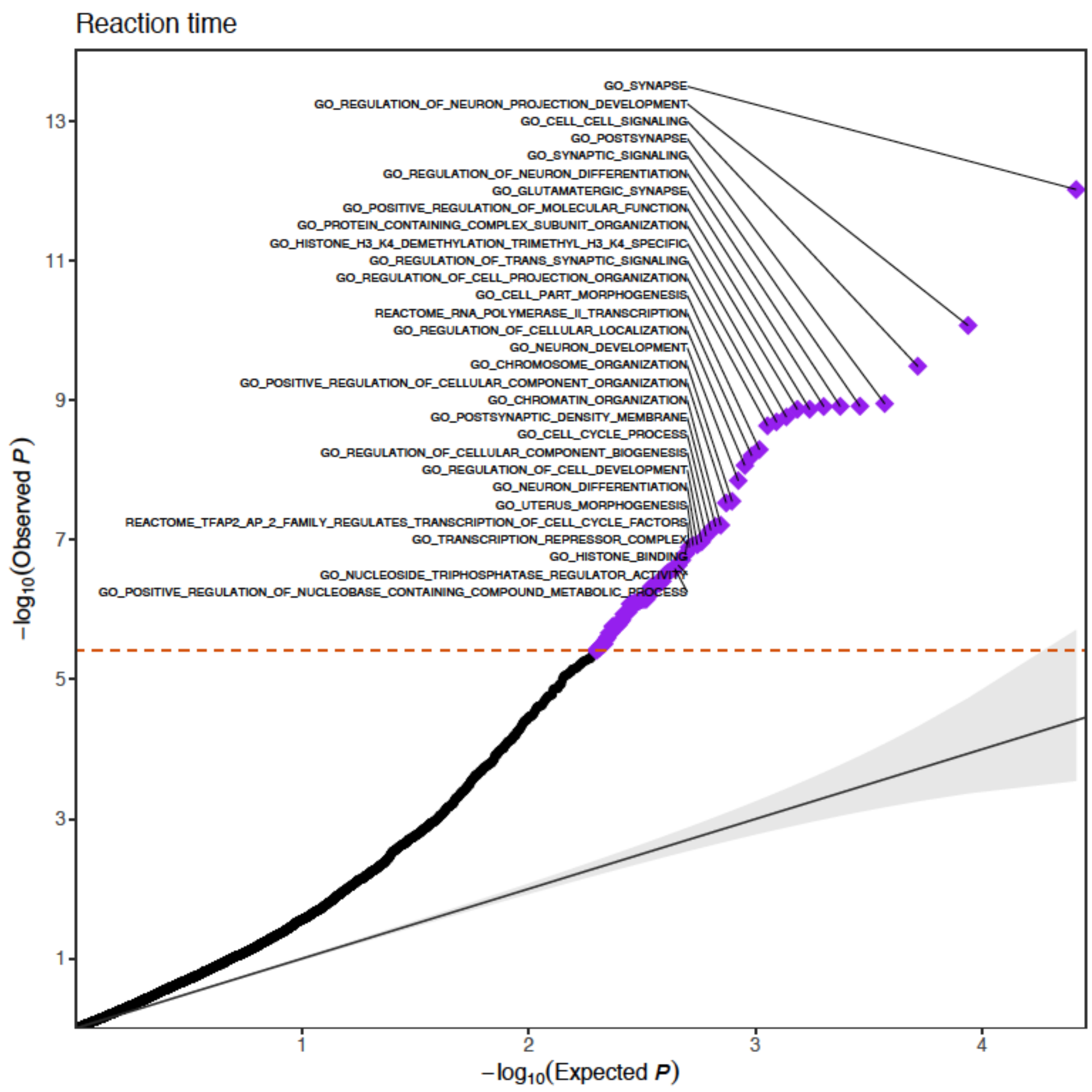

Reaction time
GO_SYNAPSE
GO_REGULATION_OF_NEURON_PROJECTION_DEVELOPMENT
GO_CELL_CELL_SIGNALING
GO_POSTSYNAPSE
GO_SYNAPTIC_SIGNALING
GO_REGULATION_OF_NEURON_DIFFERENTIATION
GO_GLUTAMATERGIC_SYNAPSE
GO_POSITIVE_REGULATION_OF_MOLECULAR_FUNCTION
GO_PROTEIN_CONTAINING_COMPLEX_SUBUNIT_ORGANIZATION
GO_HISTONE_H3_K4_DEMETHYLATION_TRIMETHYL_H3_K4_SPECIFIC
GO_REGULATION_OF_TRANS_SYNAPTIC_SIGNALING
GO_REGULATION_OF_CELL_PROJECTION_ORGANIZATION
GO_CELL_PART_MORPHOGENESIS
REACTOME_RNA_POLYMERASE_II_TRANSCRIPTION
GO_REGULATION_OF_CELLULAR_LOCALIZATION
GO_NEURON_DEVELOPMENT
GO_CHROMOSOME_ORGANIZATION
GO_POSITIVE_REGULATION_OF_CELLULAR_COMPONENT_ORGANIZATION
GO_CHROMATIN_ORGANIZATION
GO_POSTSYNAPTIC_DENSITY_MEMBRANE
GO_CELL_CYCLE_PROCESS
GO_REGULATION_OF_CELLULAR_COMPONENT_BIOGENESIS
GO_REGULATION_OF_CELL_DEVELOPMENT
GO_NEURON_DIFFERENTIATION
GO_UTERUS_MORPHOGENESIS
REACTOME_TFAP2_AP_2_FAMILY_REGULATES_TRANSCRIPTION_OF_CELL_CYCLE_FACTORS
GO_TRANSCRIPTION_REPRESSOR_COMPLEX
GO_HISTONE_BINDING
GO_NUCLEOSIDE_TRIPHOSPHATASE_REGULATOR_ACTIVITY
GO_POSITIVE_REGULATION_OF_NUCLEOBASE_CONTAINING_COMPOUND_METABOLIC_PROCESS
13
11
9
7
5
3
1
1
2
3
4
−log10(Observed P)
−log10(Expected P)

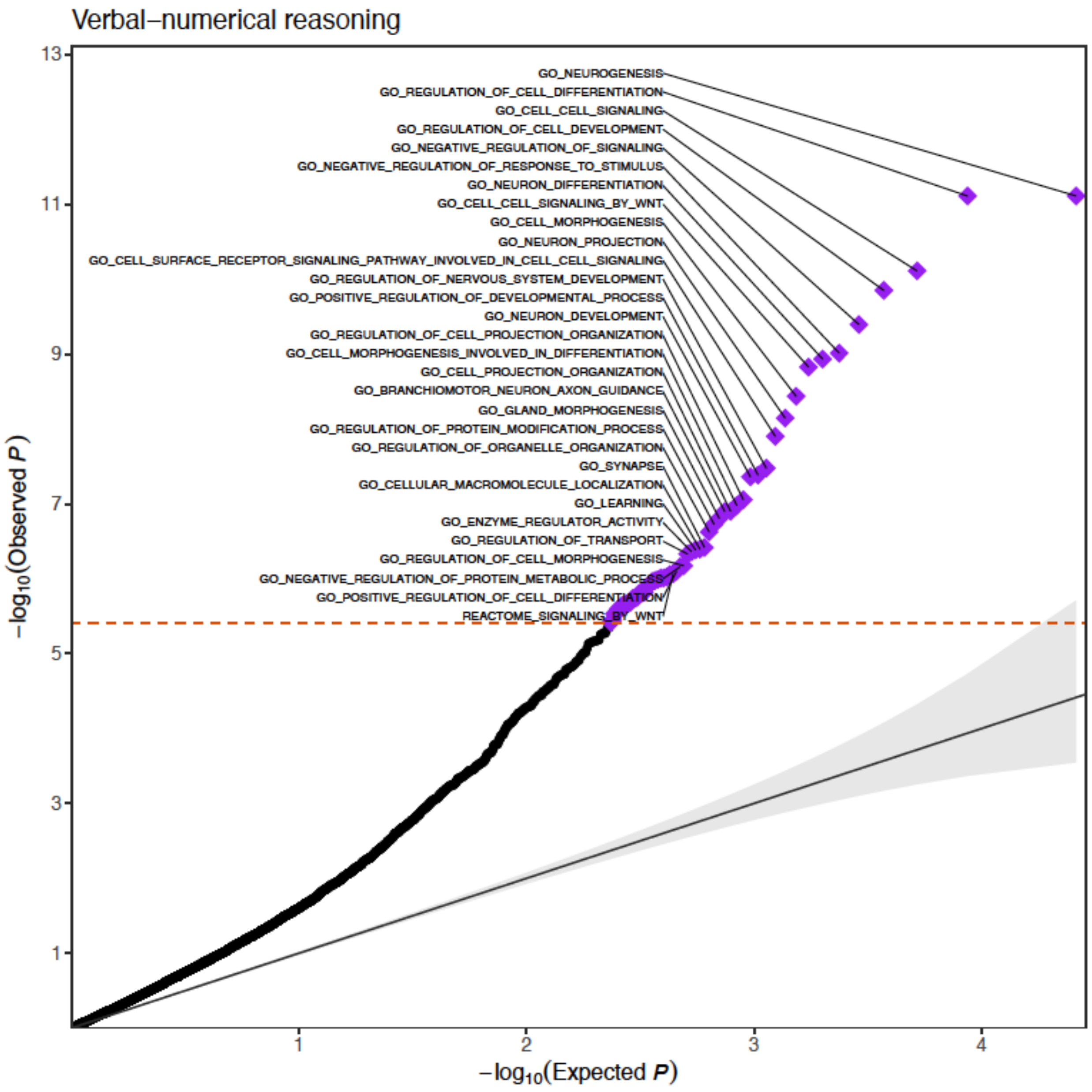


**Fig. S16. Gene set-based PTV burden analysis in European samples in the UK Biobank for educational attainment, Reaction time and verbal-numerical reasoning.** Top 30 gene sets were labeled in the figure. A total of 13,011 gene set from MSigDB v7.2 were identified, including C2 canonical pathways (N=2,808) and C5 Gene Ontology biological process (N = 7,531), cellular component (N = 996), and molecular function (N = 1,676).

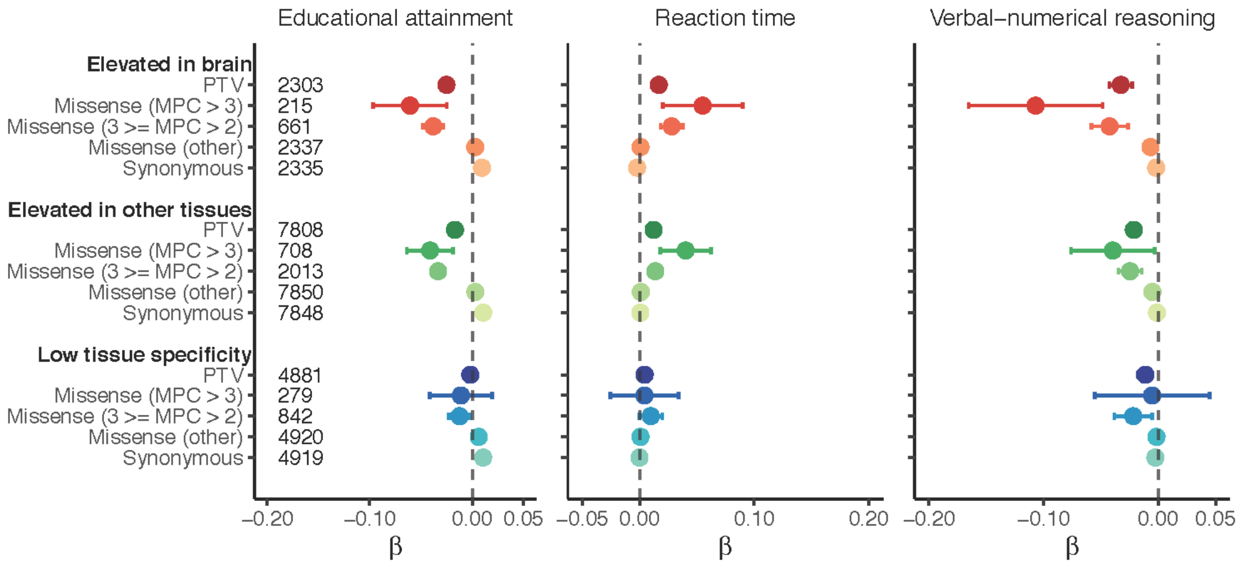


**Fig. S17. The effects of protein-truncating, missense (stratified by MPC) and synonymous variant burden in genes stratified by brain-specific expression.** Genes were stratified by elevated expression in brain tissue (2,587 genes), elevated expression in other tissues but also expressed in brain (5,298 genes) and no tissue specific expression (8,342 genes). Number of genes included in the burden is annotated for each set.

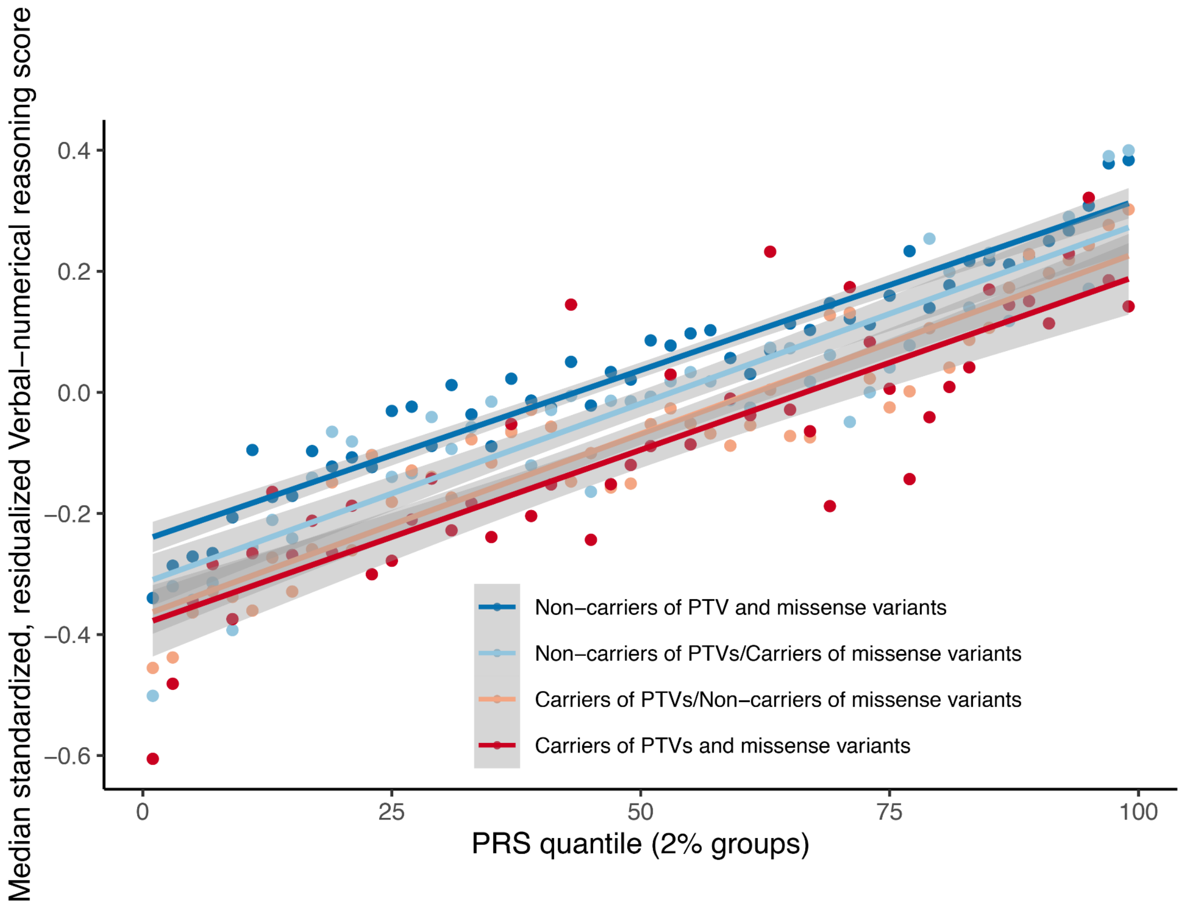


**Fig. S18. The impact of cognitive function polygenic score and carrier status of PTV and/or damaging missense variants (MPC>2) in LoF intolerant genes (pLI>0.9) on verbal-numerical reasoning.**
